# Phenotypic and genetic factors are associated with absence of cardiomyopathy symptoms in PLN c.40_42delAGA carriers

**DOI:** 10.1101/2022.05.12.22274763

**Authors:** Esteban A. Lopera-Maya, Shuang Li, Remco de Brouwer, Ilja M. Nolte, Justin van Breen, The Netherlands ACM/PLN registry, Lifelines Cohort study, Jan D.H. Jongbloed, Morris A. Swertz, Harold Snieder, Lude Franke, Cisca Wijmenga, Rudolf A. de Boer, Patrick Deelen, Paul A. van der Zwaag, Serena Sanna

## Abstract

The c.40_42delAGA variant in the phospholamban gene (*PLN*) has been associated with dilated and arrhythmogenic cardiomyopathy, with up to 70% of carriers experiencing a major cardiac event by age 70. However, other carriers remain asymptomatic or show only mild symptoms in old age. To understand the mechanisms behind this incomplete penetrance, we evaluated potential phenotypic and genetic modifiers in 74 PLN c.40_42delAGA carriers identified in 36,339 participants of the Lifelines population cohort. Asymptomatic carriers (N=48) showed shorter QRS duration (-5.73 ms, p-value=0.001) compared to asymptomatic non-carriers and symptomatic carriers (N=26), and we replicated this in different subset of 21,771 participants from the Lifelines cohort (-3.87 ms, p-value=0.028) and in 592 carriers from the Arrhythmogenic Cardiomyopathy (ACM) PLN patient registry (-6.91 ms, p-value=0.0002). Furthermore, symptomatic carriers showed a higher correlation between genetic predisposition to higher QRS duration (PGS_QRS_) and QRS (p-value=1.98×10^-8^), suggesting that symptomatic PLN c.40_42delAGA carriers may have an increased sensitivity to the effect of genetic variation in cardiac rhythm. Our results may help improve risk prediction models for cardiac outcomes for future studies, while our approach could guide studies on genetic diseases with incomplete penetrance.

## Introduction

Inherited cardiomyopathies are genetic disorders that cause cellular and/or molecular imbalances that affect the functioning of the cardiac muscle. Different cardiomyopathy subtypes now recognized: hypertrophic cardiomyopathy (HCM), dilated cardiomyopathy (DCM), arrhythmogenic cardiomyopathy (ACM), left ventricular non-compaction cardiomyopathy and restrictive cardiomyopathy.^1^ Cardiomyopathy symptoms can vary from mild symptoms to life threatening arrhythmias or end-stage heart failure (HF), and the absence or presence of a phenotype is established using a mix of imaging and electrocardiographic studies. More than 60 genes have now been associated with one or more inherited forms of cardiomyopathy, with all forms characterized by incomplete penetrance and variable expression.^2^

The gene encoding phospholamban (PLN) is one of these cardiomyopathy-associated genes. PLN is a regulator of the sarcoplasmic reticulum Ca^2+^ (SERCA2a) pump and has been implicated in Ca^2+^ homeostasis in cardiac muscle cells.^3^ Several variants in the *PLN* gene have been associated with HF.^4^ Previous studies identified a founder effect for one of these variants, the c.40-42delAGA (p.Arg14del) pathogenic variant (ClinVar accession number VCV000044580.20), in the Netherlands, particularly in the northern part of the country,^5^ that has resulted in a relatively large number of carriers in the Dutch population (estimated allele frequency 0.07%–0.22%). This variant has been associated with both DCM and ACM and is characterized by an increased risk of developing malignant ventricular arrhythmias (VAs) and end-stage HF, with age-dependent incomplete penetrance (>50% of carriers show symptoms above the age of 60 years).^6–8^

Given the high penetrance of *PLN* c.40-42delAGA, carriers at risk or subjects from suspected families are screened by a cardiologist for early disease detection. However, the under-explored mechanisms behind the incomplete penetrance of this variant create challenges in determining risk for carriers and thus hamper the decision-making process for therapies. To understand the incomplete penetrance and to support early disease detection, researchers have been looking for environmental or genetic factors that modify the risk of disease manifestation. Typically, this research only collects and analyzes phenotypical, environmental and demographic information for carriers.^8–12^ One very recent study by Verstraelen et al^13^ used abnormalities observed in carriers to calculate the risk for malignant VA and guide the decision of when to start or withhold preventive therapies, including intracardiac defibrillators. Moreover, the search for risk predictors has been extended to investigate the cumulative impact of common genetic variations previously associated to cardiac-related traits in the form of polygenic scores (PGSs), an approach that has generated preliminary results for both HCM and DCM.^14–16^

While these approaches have provided encouraging evidence that phenotypic and genetic modifiers may contribute to the observed variability in disease penetrance and expression, they cannot explain it in full. The main limitation of these studies is that they completely leave out information from the general population and focus only on factors that increase the risk of developing cardiomyopathy symptoms (i.e. differences between symptomatic and healthy carriers), an approach that could miss potential protective factors that prevent carriers from developing symptoms (i.e. differences of the healthy carriers with symptomatic carriers and general population). An exemplary scenario of the potential gain of including the general population in these designs comes from studies in sickle cell and thalassemia patients, where individuals with milder symptoms showed a higher frequency of common genetic variants in the *BCL11A* gene, which had previously been associated with increased production of fetal hemoglobin, as compared to both patients with severe symptoms and to the general population.^17,18^ It is only recently that large-scale general population biobanks have enabled unbiased identification of the *PLN* c.40-42delAGA variant, providing more power to detect potential protective factors compared to a patient cohort. To our knowledge, no such analysis has been conducted for the detection of potential protective factors to explain the incomplete penetrance of *PLN* c.40-42delAGA in cardiomyopathy.

In this study, we aimed to fill this gap by performing association analysis between the absence of cardiomyopathy symptoms and genetic and phenotypic information available for 36,339 volunteers of the Lifelines cohort, a population biobank from the Northern Netherlands.^19^ In the absence of the imaging-based measurements typically used for the diagnosis of cardiomyopathy, such as chest X-ray and echocardiogram, we defined the symptomatic group using a combination of electrocardiographic (ECG) abnormalities suggestive of *PLN*-associated cardiomyopathy and self-reported heart disease symptoms at baseline assessment or during the 5-year follow-up visit (**Methods**). This identified 74 carriers, and we defined the 48 of them who did not report or show signs or symptoms as asymptomatic. We then compared the distribution of quantitative phenotypes and genetic variation in asymptomatic carriers to the general population (asymptomatic non-carriers) (**Figure 1**). We found that, in asymptomatic carriers, QRS duration and heart rate (HR) variability were significantly lower, while HR was increased. We then replicated the lower QRS duration that we observed in asymptomatic carriers in two additional cohorts, providing the first robust evidence for factors that associate with absence of symptoms in carriers. We also observed higher correlation between the PGS of QRS (PGS_QRS_) and QRS in the symptomatic carriers (meaning a higher sensitivity for common variants effect in symptomatic carriers). While not enough genetic information was available in the replication cohorts to confirm these findings, they do support emerging evidence of the polygenic nature of many Mendelian disorders.^14,15,20,21^

**Figure 1.**
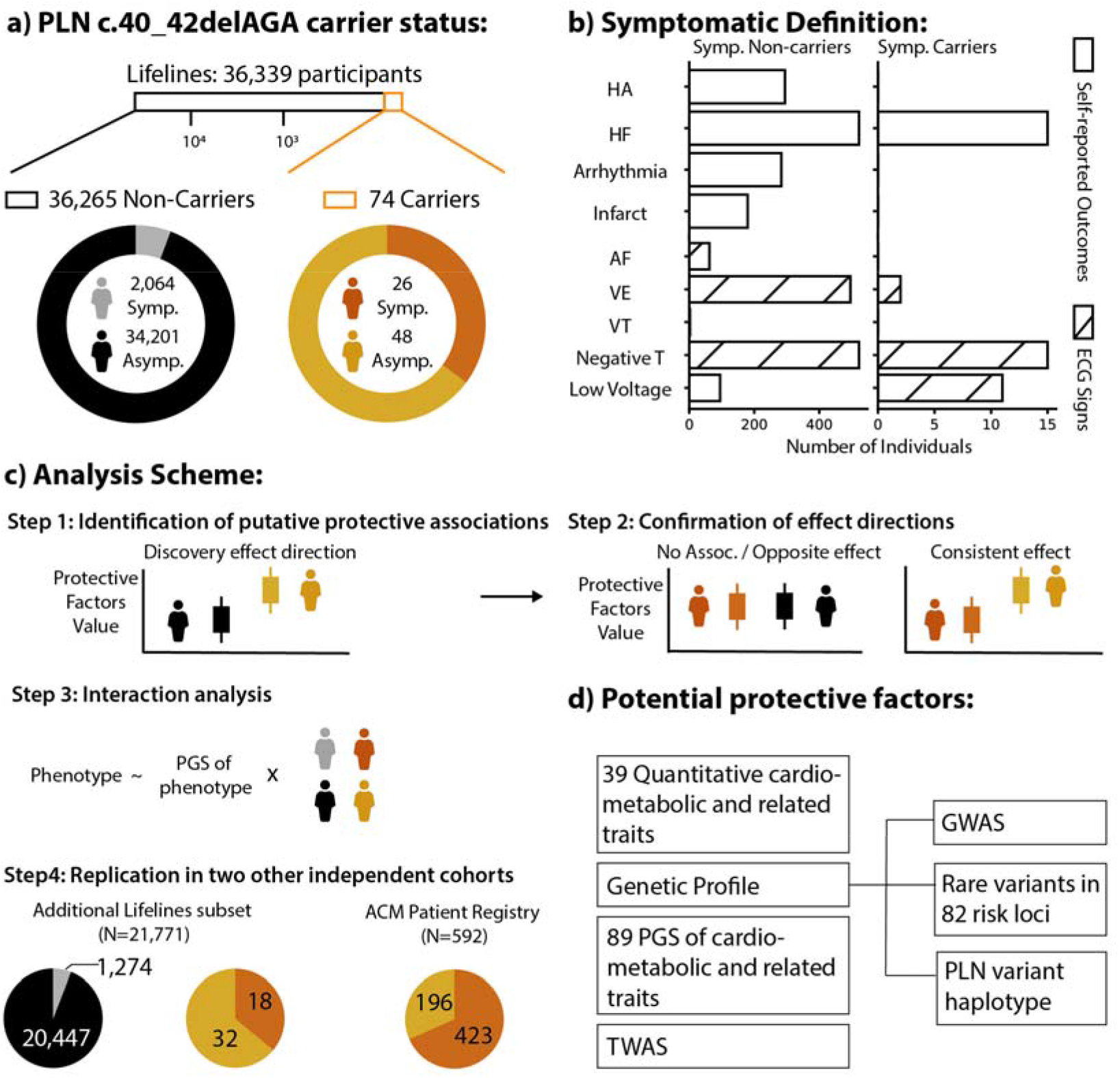
Graphic Summary. a) Distribution of the c.40_42delAGA carriers and non-carriers and ratio of the symptomatic and asymptomatic group in the subset of the Lifelines cohort. b) Definition of symptomatic status and relative abundance of people showing each clinical outcome or abnormal ECG signs. c) Analysis scheme used in this study. Briefly, we screen for potential protective factors by comparing them between asymptomatic non-carriers and asymptomatic carriers and then confirm them by comparing with the other groups (see **Methods**). Further, we assessed whether there is an interaction effect between the symptoms and carrier status and the predictability of PGS for the phenotype that shows potential protective effects. d) Potential protective factors analyzed in this study. GWAS, genome-wide association study; HA, heart attack; HF, heart failure; Infarct, infarction; AF, atrial fibrillation; TWAS, transcriptome-wide association study; VE, ventricular ectopic beats; VT, ventricular tachycardia.

## Results

### Distribution of PLN c.40_42delAGA variant and clinical profile of Lifelines samples

Genotyping data, including genotyping information for the PLN c.40_42delAGA variant, was available for 36,339 Lifelines participants with an average age of 39.9y (range 4.0–90.0y, 58.5% males and 41.5% females). Among them, we identified 74 carriers of the deletion (0.2%; 43 females and 31 males), with ages ranging from 8.0y to 68.0y and an average age of 40.9y (**Supplementary Figure 1a**). The PLN c.40_42 deletion allele showed a minor allele frequency (MAF) of 0.1%, and none of the participants were homozygous carriers of the variant. We then classified all volunteers into four groups according to the presence or absence of self-reported symptoms or ECG abnormalities suggestive of cardiomyopathy (see **Methods**) and the presence or absence of the PLN c.40_42delAGA variant. This identified 34,201 asymptomatic non-carriers, 2,064 symptomatic non-carriers, 48 asymptomatic carriers and 26 symptomatic carriers. A description of the self-reported symptoms, diseases and measured ECG signs that we used for symptomatic definition is shown in the graphic summary (**Figure 1b, Methods**).

Using this definition, we estimated an Odds Ratio (OR) of 8.98 (CI=5.33–14.79, two-sided p-value=2.18×10^-14^) for the PLN c.40_42delAGA variant in conferring risk of presenting any of the signs and symptoms and an OR of 11.2 for HF (CI=5.31–21.64, two-sided p-value=2.05×10^-8^), strongly supporting the previously reported pathogenicity of this variant. We further noticed that asymptomatic carriers were significantly younger than asymptomatic non-carriers (average ages 34.6 and 39.1, respectively, two-sided p-value=0.037), even when restricting the analyses to participants older than 40.0y (two-sided p-value=0.001, **Supplementary Figure 1b and 1c**), and we therefore used age as a covariate in all our analyses.

Considering that the Lifelines cohort has a family design, our primary analyses also accounted for familial relationships, either using the kinship matrix or removing relatives because they can introduce spurious correlation at both phenotypic and genetic level (**Methods**). While 34 of the 74 carriers did not have other relatives carrying the variant, the remaining 40 carriers belong to 17 families with two or three carriers (**Supplementary Figure 2**).

### ECG measurements are significantly different in asymptomatic carriers compared to population controls

We analyzed the 38 cardiac, metabolic and anthropometric quantitative traits that were available for at least 25 of the asymptomatic carriers. Among these traits, four were significantly different between asymptomatic carriers and asymptomatic non-carriers (multiple testing–adjusted false discovery rate q-value≤0.05, see **Methods**) after adjusting for age, gender and kinship. Specifically, asymptomatic carriers showed a significantly faster average HR of 6.41 beats/min (95%CI=3.09–9.72, q-value=2.61×10^-4^), a shorter average QRS duration of -5.7 ms (95%CI=-9.22–-2.24, q-value=1.04×10^-3^) and reduced HR variability measured by both the natural logarithm of the root mean square of successive differences (lnRMSSDc: beta=-0.182, standard error (SE)=0.090, q-value=0.0435) and by the natural logarithm of standard deviation of the normal-to-normal intervals (lnSDNN: beta=- 0.216, SE=0.094, q-value=0.0242) (**Supplementary Table 2, Figure 2**). We then repeated the analyses for these traits using phenotypes measured in the same individuals during the first Lifelines cohort follow-up visit (Visit 2), which took place approximately 5 years after the first visit, and observed consistent effects (**Figure 2**).

**Figure 2.**
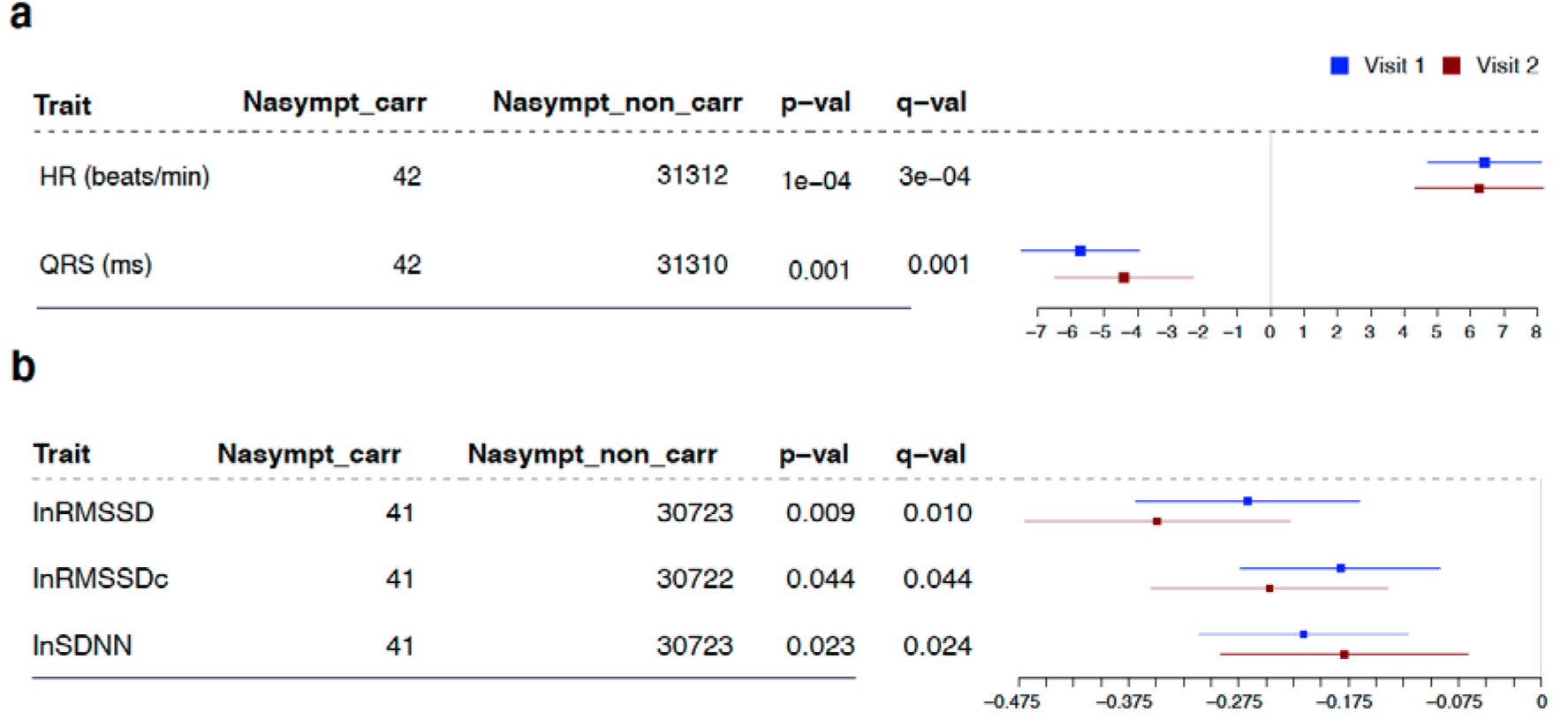
Forest plot of effect sizes for significant quantitative phenotypes. Squares represent the effect size (beta). Lines represent standard error. Blue shapes and the numbers in the columns correspond to analysis results for the baseline visit (visit 1). Red shapes correspond to the second visit approximately 5-years later (visit 2). Phenotypes are separated by the scale in a) and b) panels. Nasympt_carr, number of asymptomatic carriers with non-null information in visit 1; Nasympt_non_carr, numbers of asymptomatic non-carriers with non-null information in visit 1; p-val: p-value for the independent association; q-val: false discovery rate for conjoined analysis of all the traits that takes into account 100 permutations for each of the 38 traits; lnRMSSD: natural logarithm of the root mean square of successive differences; lnRMSSDc: RMSSD corrected by heart rate; lnSSDN: natural logarithm of the normal-to-normal intervals.

To exclude the possibility that these results were being driven by a direct impact of the PLN c.40_42delAGA variant on the phenotype, we compared the distribution of these significant phenotypes among all four groups in our analysis scheme (**Methods**). Here we observed a consistent association between high HR and low QRS duration and absence of symptoms when comparing asymptomatic carriers to symptomatic non-carriers (**Figure 3**): asymptomatic carriers showed significantly higher HR that was faster by 5.58 beats/min (95%CI=1.63–9.54, two-sided p-value=0.0057) and a QRS duration that was -7.24 ms lower (95%CI=-11.92–2.56, two-sided p-value=0.0024). Interestingly, we also found a higher QRS in symptomatic carriers (8.52 ms higher, 95%CI=4.11–12.93, two-sided p-value=1.5×10^-4^) compared to asymptomatic non-carriers, an opposing effect to that seen when comparing the asymptomatic carriers with this group. In contrast, HR variability features were not significantly different in asymptomatic carriers compared to other groups (**Supplementary Figure 3**).

**Figure 3.**
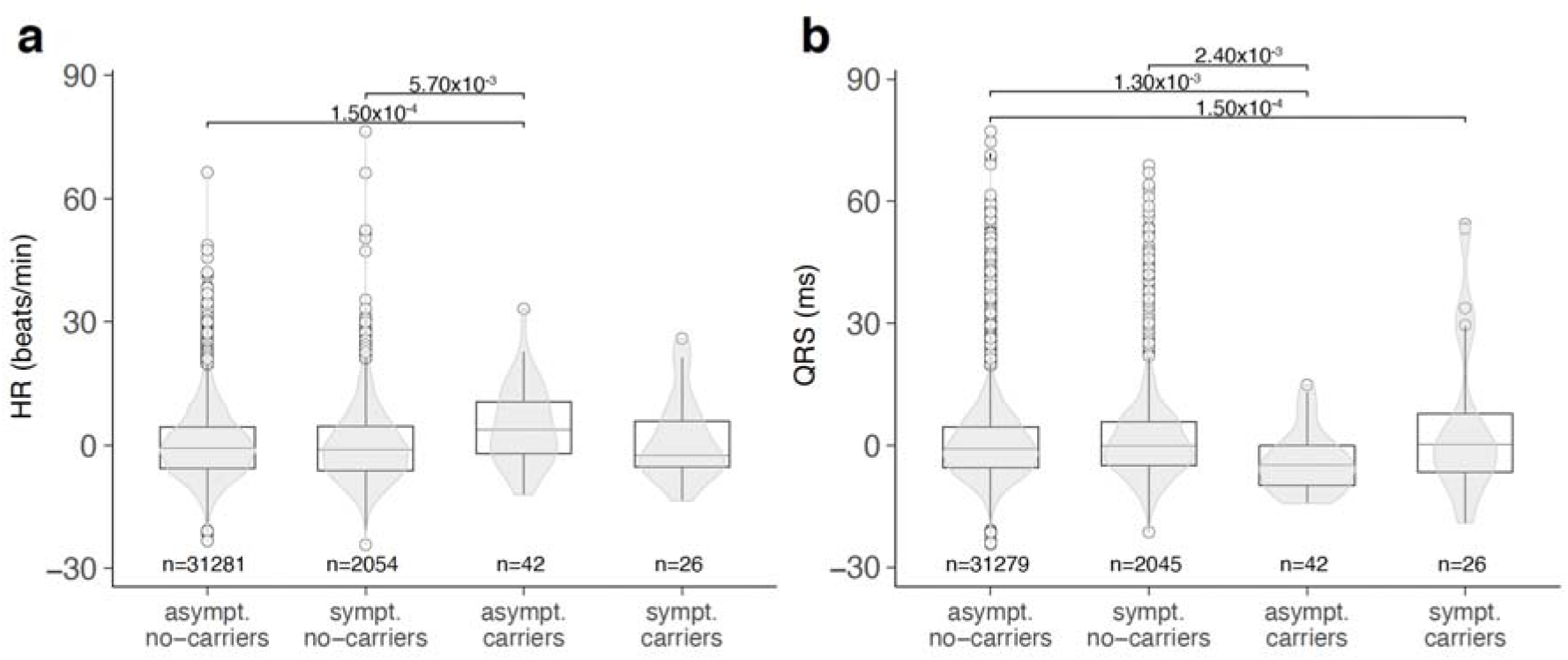
Distribution of HR and QRS with carrier status and symptom manifestation. For each phenotype, the distribution within groups is represented in two forms: the truncated violin plots in gray display the density, while the boxplots display the median (center horizontal line), the first and third quartiles (box hinges) and the values up to 1.5 interquartile ranges (IQR) away from the respective hinges (upper and lower whiskers). Values above or below these extremes are represented as individual points. a) HR. b) QRS duration. The different distribution for asymptomatic carriers compared to all the other groups is visually apparent for both these traits but is more variable in the HR variability-related traits (**Supplementary Figure 3**). P-values correspond to the regression coefficient of the adjusted trait (Y-axis) in the model described in equation 1, comparing the groups indicated by the ticks. asympt, asymptomatic; sympt, symptomatic.

### Association of shorter QRS duration with absence of symptoms in carriers replicates in an independent population cohort and a patient registry

We sought to replicate the associations we observed for HR and QRS in two additional cohorts: a second subset of the Lifelines cohort and a cohort of PLN c.40_42delAGA carriers from the ACM/PLN patient registry,^22^ which also includes asymptomatic carriers.

The Lifelines subset used for replication comprised 21,771 volunteers (62.7% females) with an average age of 41.8y (range 8.0–93.0y), of which 50 carry the PLN c.40_42delAGA variant. Following the criteria for group assignment from the primary analysis (**Figure 1**), this cohort was divided into 32 asymptomatic and 18 symptomatic carriers and 20,447 asymptomatic and 1,274 symptomatic non-carriers. When comparing the distribution of QRS and HR between the two Lifelines subsets, we observed the same effect direction and comparable magnitude (Cochran’s Q test two-sided p-value for differences in effect size=0.134) for QRS duration only (decrease=-3.87 ms, 95%CI=-7.79–0.056, one-sided p-value=0.028, **Figure 4, Supplementary Table 3A**). Since some of the volunteers in this second Lifelines subset were relatives of volunteers in our discovery set, we carried out a joint analysis, adjusting by batch-effect and relatedness, and confirmed the legitimacy of this association (p_discovery_=1×10^-3^ vs p_joint_=2×10^-4^, **Supplementary Table 3A**).

**Figure 4.**
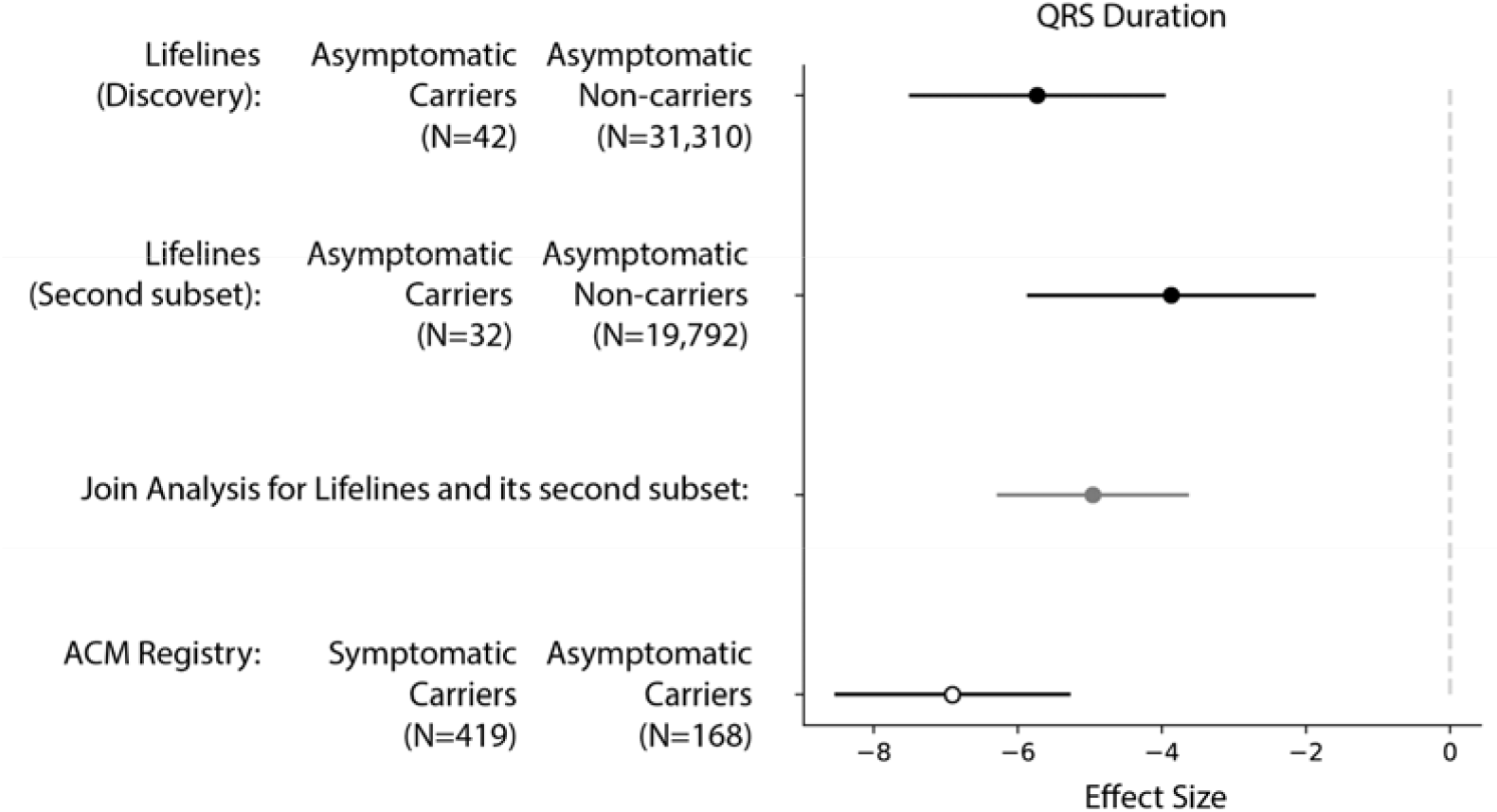
Comparison of the effect sizes for HR and QRS duration in the two replication datasets and the discovery dataset. Figure shows the effect sizes from the Lifelines discovery cohort and the two replication cohorts – an additional subset of Lifelines participants and the ACM/PLN registry. Note that the effect in the Lifelines cohorts reflects the difference between the asymptomatic carriers and asymptomatic non-carriers, whereas replication in the ACM/PLN registry compares symptomatic carriers to asymptomatic carriers.

The observation of low QRS duration was also confirmed in the ACM/PLN patient registry, which comprises 592 carriers of the PLN c.40_42delAGA variant with an average age of 41 (range 2.0–80.0y) (**Supplementary Figure 10**). Among the 592 carriers, 423 were classified as symptomatic and 169 as asymptomatic. We analyzed the heart-related phenotypes collected during each individual’s first visit using the same model used in the discovery cohort (**Methods**) except for the genetic principal component (PC) confounders, which were not available for this study because no genome-wide genotyping data was available. Here we observed consistent replication for QRS durations (**Figure 4, Supplementary Figure 10**,**Supplementary Table 3B**). Asymptomatic carriers in the ACM/PLN registry showed significantly decreased QRS duration (-6.91 ms, 95%CI=-3.69–-10.12, two-sided p-value=2.0×10^-4^) compared to symptomatic carriers, and this effect was consistent even after stratifying the cohort into younger (<40.0y) and older (≥40.0y) carriers (**Supplementary Figure 10, Supplementary Table 3B**).

### Two common genetic variants are associated with absence of symptoms in carriers

We then searched for genetic factors that could explain the incomplete penetrance of PLN c.40_42delAGA. We first analyzed the genetic region near the variant to look for variants that could explain symptoms and signs of disease manifestation in carriers. We observed that the majority of carriers (N=70) share a long identical haplotype of common and low frequency variants (MAF ≥ 0.01) of 1.38 Mb (from 604 Kb to -766 Kb from the PLN gene), which is consistent with previous observations^7^, while the remaining four samples carried distinct haplotypes varying from 1 to 330 single-nucleotide polymorphisms (SNPs). We then took a closer look by including rare variants with minor allele counts ≥ 4 in the carrier chromosomes, which allowed us to test more SNPs differentiating haplotype groups (n ≥ 3). However, neither these SNPs nor those at the boundaries of the long and shared haplotypes were significantly associated with manifestation of symptoms (**Supplementary Table** 4 and **Supplementary Figure 7**).

We next expanded our search for potentially associated genetic factors to the rest of the genome by analyzing the single additive effect of 16,478,162 common variants in autosomes (see **Methods**). For this analysis, we used only unrelated samples (39 asymptomatic carriers and 20,103 asymptomatic non-carriers). Here we observed a long stretch of hundreds of variants located on chromosome 6 spanning a region of ∼9.6 Mb that show significant associations at a classical genome-wide threshold of p-value ≤ 5×10^-8^ (**Supplementary Figure 8, Supplementary Table 5**). All these associations, however, could be attributed to common variants tagging the haplotype carrying the PLN c.40_42delAGA, and thus this association was not related to presence/absence of symptoms (**Supplementary Text**). We also found two associations independent of PLN c.40_42delAGA (**Supplementary Figure 14**) that were close to the classical genome-wide significance threshold of 5×10^-8^: one in chromosome 3 with SNP rs6768326 (allele G beta=1.52, SE= 0.29, two-sided p-value=9.26×10^-8^, **Figure 5a**) located near (∼180 Kb) the gene *RAP2B* and one in chromosome 16 with SNP rs112525682 (allele A beta=1.42, SE= 0.28, two-sided p-value=2.64×10^-7^, **Figure 5b**) located in the gene *GSE1*. However, we were unable to test for replication of these signals in the additional Lifelines subset as none of these SNPs were directly available due to the different genotyping platforms (**Methods)**. For the signal in chromosome 3, we could assess a proxy SNP, rs6791558 (r2=0.85 rs6768326 in 1000 Genomes Europeans) but found no significant association with asymptomatic carriers in the replication cohort (beta=0.27, SE=0.40, two-sided p-value=0.49, **Supplementary Table 5C**). No proxy SNP was available for the signal in chromosome 16, so this SNP could not be tested.

**Figure 5.**
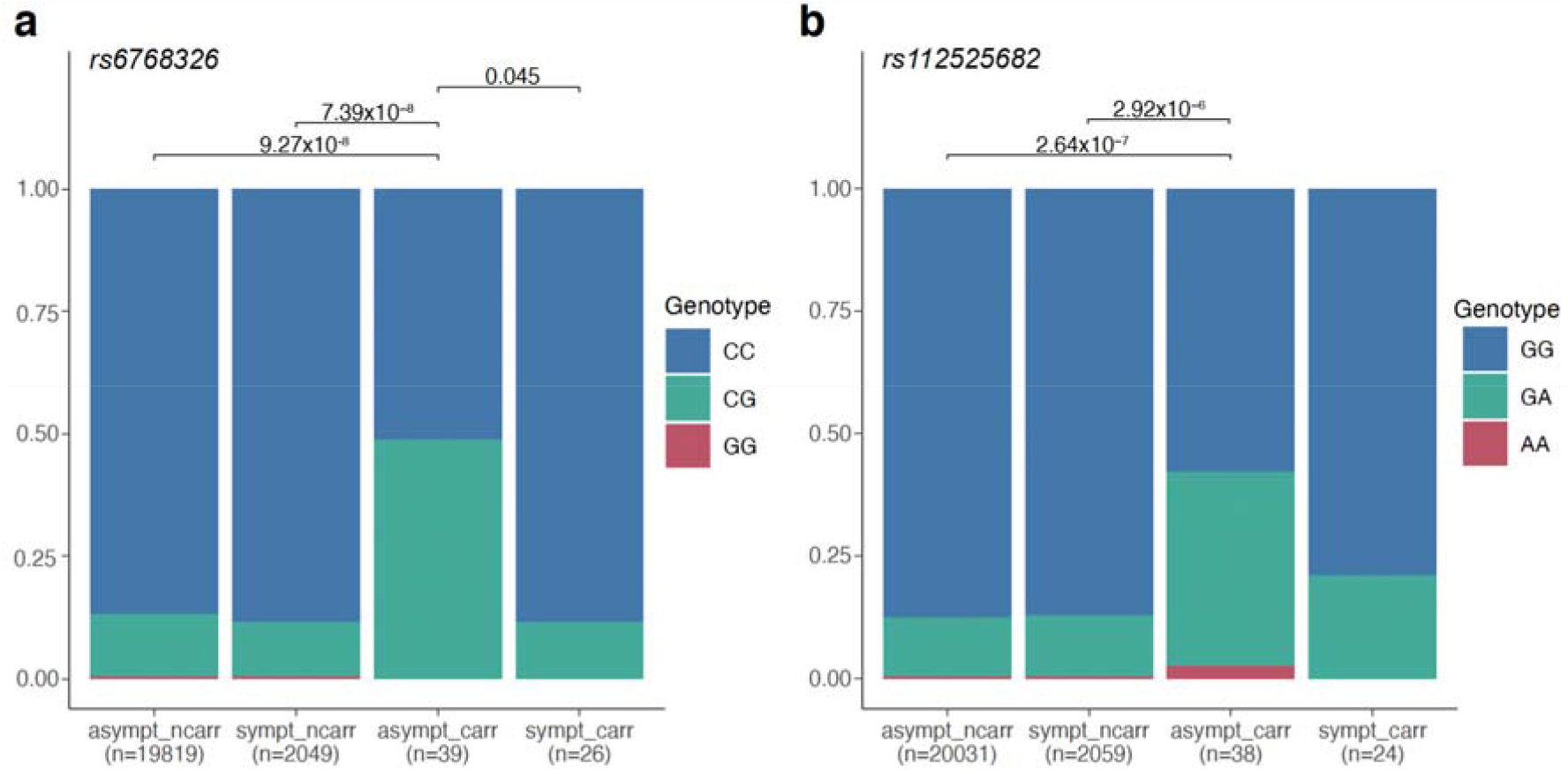
Genotype distribution of SNPs in chromosome 3 and 6 according to carrier status and symptoms manifestations. Frequency bar plots depicting the genotypes for a) SNP rs6768326 in chromosome 3 and b) SNP rs112525682 in chromosome 16. Stacked bars are used to represent the relative counts of individuals with 2 (red), 1 (green) or 0 (blue) alternative alleles. Discrete genotypes were derived by rounding the allele dosage to the nearest integer value or setting it to missing if their value was more than 0.3 away. p-values are derived from linear models adjusted for age, sex and the first PC.

### Rare variant loads in cardiac-related genes do not associate with presence or absence of symptoms in PLN c.40_42delAGA carriers

Finally, we searched for any potential aggregated effect of rare variants at 80 genomic regions previously associated with cardiomyopathies or cardiac phenotypes by GWAS and for the regions surrounding our two GWAS signals on chromosome 3 and chromosome 16 (see **Methods**). Here we observed a significant association for variants in the gene *SLC35F1* when using the low frequency threshold (MAF≤0.05, two-sided p-value=4.3×10^-22^) and the rare variant threshold (MAF≤0.01, two-sided p-value=2.8×10^-26^) for variant inclusion (**Supplementary Table** 6). Subregion analyses in the same gene revealed significant associations with asymptomatic carriers in regions containing exons 1, 2, 4 and 8. These results are interesting because the common variants near *SLC35F1* are already known to associate with QRS duration^23^, QT interval^24^ and resting HR.^25^ However, this gene is also in close proximity (<250kb) to the PLN gene and spurious association may thus arise due to different combinations of variants in the core haplotype shared by carriers compared to non-carriers. Due to the limited sample size of the symptomatic carrier group, we could not compare symptomatic carriers with asymptomatic carriers to show a different load of rare variants between groups. However, when comparing the rare variant burden in the same sub-regions between symptomatic carriers and asymptomatic non-carriers, these were also significantly different and had the same effect direction (**Supplementary Table** 6). We therefore concluded that these signals are the result of a confounding effect of the founder haplotypes of the PLN c.40_42delAGA variant and cannot be attributed to the presence or absence of symptoms.

### Polygenic burden for left ventricular end systolic volume differs between asymptomatic and symptomatic carriers of the PLN c.40_42delAGA variant

We next evaluated potential associations between common genetic variants known to associate with cardiac traits and absence of symptoms by calculating PGSs for 89 unique traits and compared PGS distributions between asymptomatic carriers and asymptomatic non-carriers (**Methods and Supplementary Table 7A**). Several PGSs were significantly different (false discovery rate q-value <0.05, see **Methods**) in asymptomatic carriers (**Supplementary Figure 4** and **Supplementary Table 7B**), but no PGSs showed significant differences when comparing symptomatic and asymptomatic carriers **(Supplementary Figure 5)**. When we repeated the associations using PGSs calculated without the variants located in chromosome 6, we only observed a lower PGS for left ventricular end systolic volume corrected by body surface area (LVESVI) in asymptomatic carriers (PGS_LVESVI_, beta=-0.30, SE=0.15, q=0.035), which indicates that the other differences can be attributed to common variations located near the PLN c.40_42delAGA variant (**Supplementary table 7E**).

### Symptomatic carriers PLN c.40_42delAGA variant are more sensitive to the polygenic burden of QRS duration

To further investigate the effect of PGSs for cardiac measurements, we modeled their prediction utility on their respective measurements and potential interactions with age, gender and participant group assignment. Here we found that the PGS_QRS_ has a stronger impact for predicting QRS in symptomatic carriers (two-sided p-value=4.27×10^-11^, **Figure 6b, Supplementary Table 8**) compared to the other groups, and this interaction remained significant when applying inverse-rank normalization to the trait (two-sided p-value=1.53×10^-5^) and when calculating PGS_QRS_ without chromosome 6, thus excluding variants within or near *PLN* (two-sided p-value=2.30×10^-5^, **Supplementary Table 8**). Furthermore, PGS_QRS_ was not associated with the severity of symptoms and signs observed in symptomatic carriers (**Supplementary Figure 6**). We also found a significant interaction for PGS_PR_, but this was not robust to normalization of the measurement.

**Figure 6.**
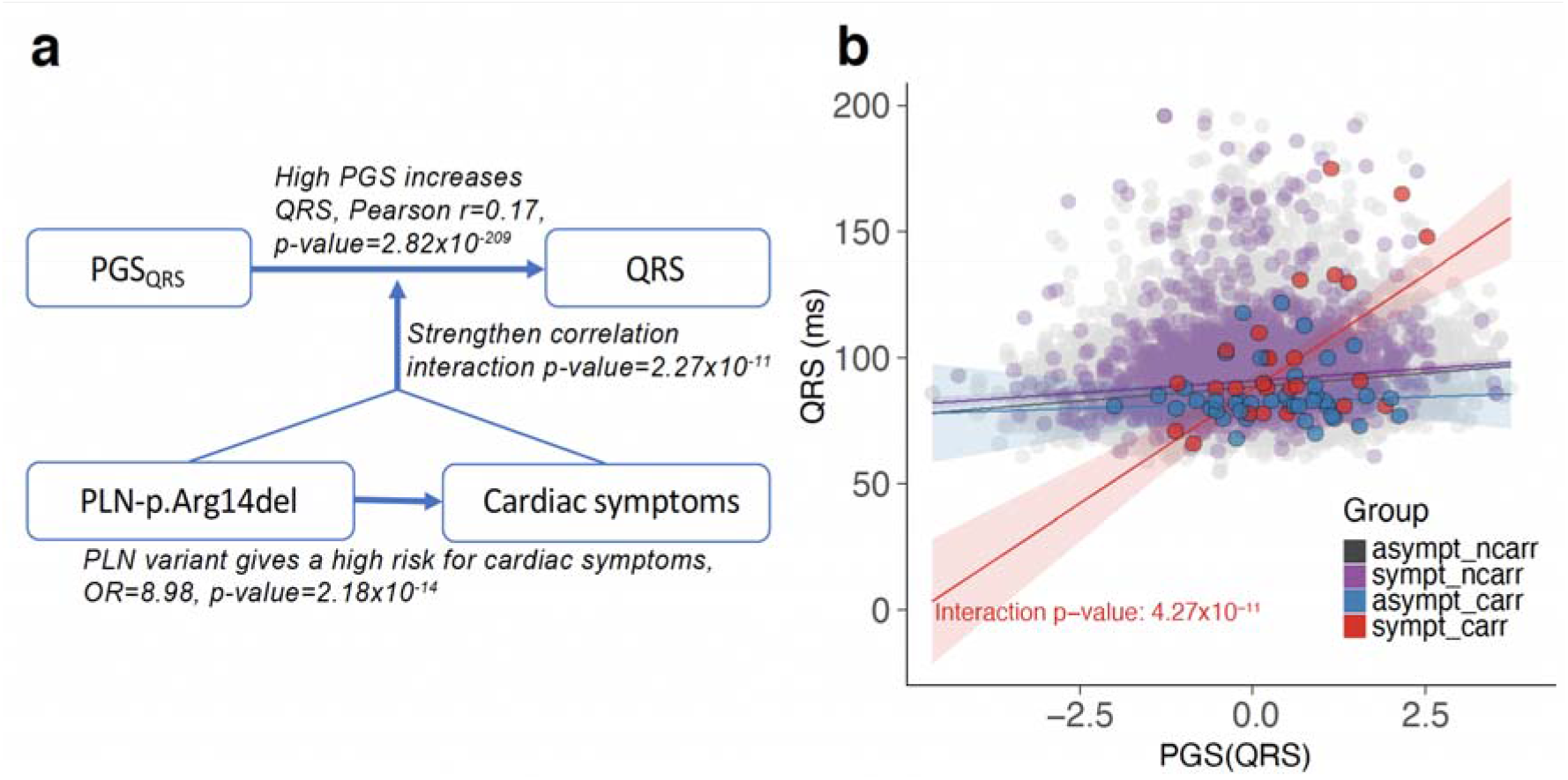
Interaction of PGS_*QRS*_ with symptomatic carrier status. a) Schematic overview of the interaction. PLN c.40_42delAGA carriers show an increased risk of developing cardiac symptoms. In turn, symptomatic carriers show a higher correlation between PGS_QRS_ and QRS compared to asymptomatic non-carriers. b) Scatter plot and regression line for the partial correlation between PGS_QRS_ and QRS for each group studied (colors according to the legend).

### No evidence for gene expression differences predicted by common variants

Finally, we searched for potential gene expression differences that could explain the presence/absence of symptoms in PLN c.40_42delAGA carriers by performing a transcriptome-wide association Study (TWAS). Briefly, we used PrediXcan^26^ to impute gene expression levels for up to 8,610 genes in the tissues artery aorta, artery tibial, heart atrial appendage and heart left ventricle and compared the imputed gene expression between asymptomatic carriers and asymptomatic non-carriers and between symptomatic carriers and symptomatic non-carriers (**Methods**). Expression of the *PLN* gene was also predicted and evaluated. Among all the genes tested, four (*MARVELD2, LAMA4, FAM184A* and *NCBP1*) were significantly different in one of the comparisons and one (*NT5DC1*) was significantly different in both comparisons. We closely examined the TWAS results in the most relevant tissue, heart left ventricle, but found that these association signals were similar in size and effect direction in both comparisons (**Supplementary Figure 9, Supplementary Table 9**), therefore differences in these genes cannot be related to presence/absence of symptoms in PLN c.40_42delAGA variant carriers.

## Discussion

In this study we used the Lifelines Dutch general population biobank to search for modifiers of the PLN c.40_42delAGA variant, a Dutch founder variant known to increase a carrier’s risk of developing malignant VA and HF. We observed that QRS duration was significantly shorter in c.40_42delAGA carriers who showed no signs or symptoms of the disease as compared to other carriers and non-carriers. We then replicated this observation in another subset of the same population and in a patient registry. QRS duration reflects ventricular activation and depolarization,^27^ and QRS widening has been established as an intrinsic element of HF that comes with worse outcomes. In line with this, deformations of the QRS complex are frequently found in DCM patients,^28^ and an increased QRS (≥120ms) duration is frequently observed in severe cardiac outcome (up to 47% of patients with HF).^29^ Increased QRS duration was also previously observed in symptomatic PLN c.40_42delAGA carriers compared to asymptomatic carriers,^13^ and thus, until now, increased QRS duration was only considered a risk factor. Here, however, we show that QRS duration was significantly lower in asymptomatic carriers even when compared to the general population, which suggests that short QRS may offset or postpone disease. On the other hand, QRS widening is a risk factor, as explained by its established relationship with myocardial scarring and remodeling. Intriguingly, the lower QRS duration in asymptomatic carriers and the higher QRS in symptomatic carriers could not be explained by differences in genetic predisposition conferred by common variants that were previously associated to QRS duration (PGS_QRS_).

We observe that the effect of common genetic variation on QRS duration is more pronounced in symptomatic carriers, with the correlation between PGS_QRS_ and measured QRS duration significantly stronger in symptomatic carriers of the PLN c.40_42delAGA variant. This is more complex than just a genetic interaction as PLN c.40_42delAGA carriers who are asymptomatic do not show the same aberrant sensitivity to the PGS. This effect is also unlikely to be caused by HF by itself because people with the wildtype PLN allele (non-carriers) and HF also show normal sensitivity to the PGS_QRS_. That said, we have not identified a mechanistic explanation and these results require replication in an independent cohort. However, replication will be challenging because the PLN c.40_42delAGA variant is less frequent in other populations. According to gnomAD, the variant was identified only in two individuals out of 25,328 North-western European samples sequenced, and none in the 39,118 samples of other European or non-European origin.

For now, we can only speculate as to why we observe this complex interaction effect. In this manuscript, we explored the modifiers that make the PLN c.40_42delAGA variant pathogenic in some people but not in others. However, it might also be that the genetic or environmental modifiers that influence this sensitivity to the *PLN* variant also increase the sensitivity to PGS_QRS._ This is not unlikely as the main mechanism disturbed by the PLN mutation is calcium homeostasis, we can infer that part of the pathogenicity might be explained by disturbance in this system. Several genes implicated by the GWAS on QRS are calcium-handling proteins, ^30^ and it has been shown that calcium levels can influence ventricular repolarization, although the impact on QRS duration is weak^31^.

Further studies are needed to investigate this hypothesis. For example, as larger GWAS on QRS are carried out and more associated genetic variants discovered, the increased power and resolution of PGS_QRS_ could help clarify the signal in both groups of carriers by pinpointing a subset(s) of genes involved in specific pathways through pathway-based PGS^32,33^. Studies aiming to investigate differences in non-genetic factors between symptomatic and asymptomatic carriers will be more difficult to establish. Although genetics explains less than half of the intra-individual variation in QRS,^34^ the environmental factors that contribute to differences in QRS duration in the general population are largely unknown. We can speculate that physical activity can be one of these, as athletes and those engaging in regular physical training have relevant cardiac structural and electrical changes.^35^ Still, in our Lifelines population cohort, we did not find significant associations between QRS duration and exercise or between QRS duration and participants living in less urban locations (**Supplementary Text, Supplementary Table 11A**).

We cannot rule out that the association of QRS duration that we observe is merely a proxy for other potentially protective factors that were not measured in Lifelines participants. For example, a recent study carried out on the ACM/PLN patient registry identified certain ECG- or MRI-derived features, such as left ventricular ejection fraction (LVEF) and 24-h premature ventricular contraction (PVC) count, as stronger risk factors for developing ventricular arrhythmia, as compared to QRS duration^13^. Given the existing correlation between QRS and LVEF and the 24-h PVC count (**Supplementary Figure 11, Supplementary Text**), we cannot exclude that our signal based on QRS actually captures the effect of unmeasured LVEF and 24-h PVC count in our cohort. However, genetics and experimental model studies suggest that QRS duration could have a non-null causal role in symptom manifestation. Previous studies have shown computational evidence and suggestive experimental evidence in other organisms. In Zebrafish, the *plna* R14del variant causes beat-to-beat variations in cardiac output in the absence of cardiac remodeling, suggesting that the cardiac contractile dysfunction is not caused by but rather causal for cardiac remodeling.^36^ Causal relationships between anomalies in the QRS complex, specifically in the Q-R upslope, and increased risk of DCM have been found through the Mendelian Randomization approach using genetic variants of ECG signatures^37^.

While Verstraelen et al.^13^ found QRS duration to have limited predictive ability for VA compared to other cardiac measurements, we found that its effect still significantly improved the baseline prediction model from an LOO-AUC of 0.66 to 0.72 (DeLong test p-value=0.007, **Supplementary Figure 12, Supplementary Text**). Compared to the final model using all four features (number of negative T waves and presence of low-voltage ECG, PVC count/24 h and LVEF), the baseline model excluded PVC counts and LVEF because these two features correlate highly with QRS duration (See details in **Supplementary Text**). Therefore, we suggest that QRS duration could be further explored for its clinical value in predicting other severe events and interim outcomes for PLN c.40_42delAGA carriers.

While we used a large population cohort to search for genetic variants that could act as potential modifiers, we acknowledge that it was still underpowered for finding associations at the genome-wide level (0.7% of power to detect an OR of 5.0 for a SNP with MAF=0.1). We implemented additional strategies to investigate the cumulative impact of common and rare variants through PGS and the aggregated rare variants effects, but found no significant signal. We were also limited in investigating our findings in the replication datasets because of lack of genotyping information; no genotyping information was available for the ACM/PLN registry, while the information for the second Lifelines subset came from a different genotyping array and did not include imputed variants. Therefore, we could not validate one of the two suggestively significant findings in GWAS and were unable to replicate the PGS interaction in symptomatic carriers. Finally, for the general population in the Lifelines cohort, we did not have specialized cardiac phenotypes such as cardiac MRI measurements that could be better indicators of risk or protection.

In conclusion, we used a large population cohort to identify phenotypic and genetic modifiers of PLN c.40_42delAGA, a variant that leads to an inherited genetic disorder with incomplete penetrance. By taking advantage of the unbiased identification and inclusion of carriers showing no symptoms, we identified and replicated an association between shorter QRS duration and a lack of disease signs and symptoms. We envision that this approach can be applied to other highly penetrant disorders in other large population cohorts, thereby capitalizing on the hundreds of thousands or millions of participants so that rare disease-causing variants reach a substantial number of carriers with variable symptoms.

## Data Availability

All data produced in the present work are contained in the manuscript and supplementary information, and all date used to produce the results are available upon reasonable request

https://www.lifelines.nl/researcher/how-to-apply

## Acknowledgments

The authors wish to gratefully acknowledge the services of the Lifelines Cohort Study, the contributing research centers delivering data to Lifelines, and all the study participants.

The Lifelines Biobank initiative has been made possible by funding from the Dutch Ministry of Health, Welfare and Sport, the Dutch Ministry of Economic Affairs, the University Medical Center Groningen (UMCG the Netherlands), University of Groningen and the Northern Provinces of the Netherlands (Drenthe, Friesland and Groningen). This project was carried out under Lifelines project number OV18_0473. The generation and management of GWAS genotype data for the Lifelines Cohort Study is supported by the UMCG Genetics Lifelines Initiative (UGLI). UGLI is partly supported by a Spinoza Grant from NWO, awarded to Cisca Wijmenga.

We thank Raul Aguirre-Gamboa for his contribution in informatics training and initial analyses, Mathieu Plateel and Jody Geelderloos-Arends for their contribution in genotyping the Lifelines samples, Kate McIntyre for help developing the manuscript, the UMCG Genomics Coordination Center, the UG Center for Information Technology and their sponsors (BBMRI-NL and TarGet) for storage and computational infrastructure. This study was supported by UMCG HAP grant CD017.0031/ronde 2017-2/nr324 (P.vdZ), by NWO VIDI grant number 917.164.455 (M.A.S), ERC advanced grant ERC-671274 (C.W.), by the NWO gravitation grant The Netherlands Organ-on-Chip Initiative 024.003.001 (C.W.) and Spinoza award NWOSPI 92-266 (C.W.); and by Colciencias fellowship ed.783 (E.A.L.M.), by Netherlands Heart Foundation (2017-21; 2017-11; 2018-30; 2020B005) (R.A.d.B. & R.d.B); and by the European Research Council (ERC CoG 818715) (R.A.d.B. & R.d.B); and Foundation leDucq (Cure PhosphoLambaN induced Cardiomyopathy (Cure-PLaN) (R.A.d.B. & R.d.B).

## Lifelines Cohort Study-UGLI group authors

Raul Aguirre-Gamboa^1^, Patrick Deelen^1^, Lude Franke^1^, Jan A Kuivenhoven^2^, Esteban A Lopera Maya^1^, Ilja M Nolte^3^, Serena Sanna^1^, Harold Snieder^3^, Morris A Swertz^1^, Peter M. Visscher^3,4^, Judith M Vonk^3^, Cisca Wijmenga^1^

^*1*^ *University of Groningen, University Medical Center Groningen, Department of Genetics, Groningen, the Netherlands*

^*2*^ *University of Groningen, University Medical Center Groningen, Department of Pediatrics, Groningen, the Netherlands*

^*3*^ *University of Groningen, University Medical Center Groningen, Department of Epidemiology, Groningen, the Netherlands*

^*4*^ *Institute for Molecular Bioscience, The University of Queensland, Brisbane, Queensland, Australia*.

## Netherlands ACM/PLN Registry authors

Laurens P Bosman^1^, Tom E Verstraelen^2^, Freya HM van Lint^3^, Moniek GPJ Cox^4^, Judith A Groeneweg^5^, Thomas P Mast^5^, Paul A. van der Zwaag^6^, Paul GA Volders^6^, Reinder Evertz^8^, Lisa Wong^9^, Natasja MS de Groot^10^, Katja Zeppenfeld^11^, Jeroen F van der Heijden^5^, Maarten P van den Berg^4^, Arthur AM Wilde^2^, Folkert W Asselbergs^1^, Richard NW Hauer^1^, Anneline SJM te Riele^12^, J Peter van Tintelen^12^

^*1*^ *Durrer Centre for Cardiovascular Research, Netherlands Heart Institute, Utrecht, The Netherlands*.

^*2*^ *Department of Clinical and Experimental Cardiology, Amsterdam UMC, Amsterdam, The Netherlands*.

^*3*^ *Department of Clinical Genetics, Amsterdam UMC, Amsterdam, The Netherlands*.

^*4*^ *Department of Cardiology, UMC Groningen, Groningen, The Netherlands*.

^*5*^ *Department of Cardiology, UMC Utrecht, Utrecht, The Netherlands*.

^*6*^ *Department of Genetics, UMC Groningen, Groningen, The Netherlands*.

^*7*^ *Department of Cardiology, Maastricht UMC, Maastricht, The Netherlands*.

^*8*^ *Department of Cardiology, Radboud MC, Nijmegen, The Netherlands*.

^*9*^ *Department of Cardiology, Amsterdam UMC, Amsterdam, The Netherlands*.

^*10*^ *Department of Cardiology, Erasmus MC, Rotterdam, The Netherlands*.

^*11*^ *Department of Cardiology, LUMC, Leiden, The Netherlands*.

^*12*^ *Durrer Centre for Cardiovascular Research, Netherlands Heart Institute, Utrecht, The Netherlands*.

## Netherlands ACM/PLN Registry collaborators

A. Amin (Amsterdam UMC, Amsterdam, the Netherlands); F.W. Asselbergs (UMC Utrecht, Utrecht, the Netherlands); D.Q.C.M. Barge-Schaapveld (LUMC, Leiden, The Netherlands); M.P. van den Berg (UMC Groningen, Groningen, the Netherlands); S.M. Boekholdt (Amsterdam UMC, Amsterdam, The Netherlands); L.P. Bosman (UMC Utrecht, Utrecht, the Netherlands); M.G.P.J. Cox (UMC Groningen, Groningen, the Netherlands); D. Dooijes (UMC Utrecht, Utrecht, The Netherlands); R.N.W. Hauer (Netherlands Heart Institute); A.C. Houweling (Amsterdam UMC, Amsterdam, the Netherlands); N.M.S. de Groot (Erasmus MC, Rotterdam, the Netherlands); A.S.J.M. te Riele (UMC Utrecht, Utrecht, the Netherlands); J.P. van Tintelen (UMC Utrecht, Utrecht, the Netherlands); T.E. Verstraelen (Amsterdam UMC, Amsterdam, the Netherlands); P.G.A. Volders (MUMC+, Maastricht, the Netherlands); A.A.M. Wilde (Amsterdam UMC, Amsterdam, the Netherlands); S.C. Yap (Erasmus MC, Rotterdam, The Netherlands); K. Zeppenfeld (LUMC, Leiden, the Netherlands)

## Authors contribution

Conceptualization: S.S, P.v.d.Z., J.D.H.J.

Data curation: E.A.L.M., S.L., J.B., R.B.

Formal Analysis: E.A.L.M., S.L.

Funding acquisition: S.S, P.v.d.Z., C.W., L.F., M.A.S, R.A.d.B.

Investigation: S.S., S.L., P.D., P.v.d.Z., E.A.L.M.

Methodology: S.S., S.L., P.D., E.A.L.M.

Project administration: S.S, P.v.d.Z.

Resources: H.S., R. Brouwer, R.A.d.B, M.A.S.,

ACM Registry, Lifelines Cohort Study

Software: S.L., E.A.L.M, R.B.

Supervision: S.S, P.v.d.Z., P.D., R.A.d.B, M.A.S.

Validation: E.A.L.M., S.L., I.N., J.B., H.S., R. Brouwer, R.A.d.B

Visualization: E.A.L.M., S.L., S.S, P.v.d.Z., P.D.

Writing – original draft: E.A.L.M., S.L., S.S, P.v.d.Z., P.D.

Writing – review & editing: All authors

Roles were defined by CRediT (Contributor Roles Taxonomy)

## Methods

### Cohort description

Lifelines is a multi-disciplinary population-based cohort study with a three-generation design that examines the health and health-related behaviors of 167,729 people living in the north of the Netherlands. Lifelines employs a broad range of investigative procedures to assess the biomedical, socio-demographic, behavioral, physical and psychological factors that contribute to the health, disease and quantitative traits of the general population, with a special focus on multi-morbidity and genetics.^19^ Participants visited Lifelines research sites for a physical examination, including blood pressure and ECG measurements, and blood and urine collection to measure several disease biomarkers. Data from one follow-up visit 5 years after the baseline recruitment visit is now available for most participants. The Lifelines study was approved by the medical ethical committee from the University Medical Center Groningen (METc number: 2007/152). An informed consent was collected for all participants.

### Genotype data

A subset of 38,030 Lifelines participants were genotyped within the UMCG Genotyping Lifelines Initiative (UGLI) using the Infinium Global Screening array® (GSA) Multiethnic Diseases version, which includes around 700K variants. Samples were processed at the Rotterdam Genotyping Center and the Department of Genetics of the UMCG, following manufacturer instructions. Standard quality control procedures were used to flag and remove low quality samples and genetic markers, and the genetic content of the final quality-controlled dataset was augmented through genotype imputation carried out using the Haplotype Reference Consortium panel v1.172, as described previously.^38^ Quality-controlled, genotyped and imputed genotype information was obtained for 36,339 participants and 39,131,578 genetic variants on autosomes, which were used for genetic analyses. Based on a PC analysis that analyzed all the 36,339 Lifelines participants together with the 1000 Genomes samples,^38^ only 34 participants were of non-European origin and were not used for the analyses.

Deletion/insertion of rs397516784-AGA (PLN c.40_42delAGA) was genotyped directly. Genotype intensities were screened and indicated lack of spurious genotype calls (**Supplementary Figure 13**). Quality control parameters indicated that the variant was genotyped with high quality (genotyping call rate=99.96%, Hardy-Weinberg equilibrium p-value=1). Genotypes at this variant were used to classify samples into carriers and non-carriers according to the presence or absence of the deletion. All identified carriers were of European ancestry.

### Definition of asymptomatic participants in the Lifelines cohort

We defined a participant to be asymptomatic for cardiomyopathic disease if they did not report any cardiac symptoms in the self-reported questionnaire at baseline or follow-up and the cardiologist did not report any sign of the disease during the ECG measurements at these time points. We considered the following to be symptoms of disease: HF, heart attack, infarct and being treated for arrhythmia. We defined signs of disease as any of the following indicators noted by the ECG software and reviewed by the cardiologists: low voltage or micro-voltages, negative T, ventricular tachycardia, ventricular extrasystole or atrial fibrillation. A participant presenting one or more of these symptoms or signs was classified as symptomatic.

### Analysis scheme

To identify potentially protective factors, we analyzed phenotype and genetic variables using a 2-step procedure to compare different groups of participants according to carrier status and symptom manifestation and followed up significant results with more in-depth analyses (step 3) and replication (step 4) (**Figure 1**). Specifically, in step 1 we screened all available variables by comparing asymptomatic carriers with asymptomatic non-carriers. In step 2, variables showing significant differences (q<0.05) were further investigated by comparing their distribution among the other groups. Specifically, we compared these variables between symptomatic carriers and asymptomatic non-carriers, where we expect to find either no difference or a difference in the opposite direction to that observed in step 1. At the same time, we compared these variables between symptomatic carriers and asymptomatic carriers, where we expect to find a difference in the same direction as in step 1. For these comparisons, we used the equations described in the sections below by coding the *outcome* variable accordingly.

We used permutations to derive a proper significance threshold for step 1 that accounts for the high correlation between variables and the multiple tests performed. Specifically, we derived empirical q-values from the p-values of the joint results for all variables of 1000 permutations for each variable. In each permutation, the variable’s value was shuffled independently of the genetically related variables (kinship, PCs and carrier status), which remained linked. This was applied to analyses that investigated quantitative phenotypes and PGSs. For GWAS and rare variants burden analyses, we used the standard thresholds of p<5×10^-8^ and 2.5×10^-6 39^, respectively.

Finally, to test the robustness of our results, we repeated the analyses with the normalized phenotypes using the Rank.norm() function of the RNOmni v1.0 R package,^40^ implemented in R v4.0.3.

### Quantitative phenotypes: definition and analysis

We considered 64 quantitative phenotypes representing anthropometric measures and cardiometabolic functions as potentially protective factors. These phenotypes were measured as previously described.^19^ Outliers and unrealistic values were removed, as described in our previous work,^38^ and further extreme or unrealistic values (HR>200, QRS>200, QTC>600, PQ>320) were removed for cardiac traits.^41^ We also included four quantitative traits associated with HR variability that were derived from the ECG and calculated in the Lifelines cohort in a previous work.^42^ The full description of the quantitative traits investigated included, along with cohort descriptive statistics, can be found in **Supplementary Table 1**. We decided not to use phenotypes with fewer than 25 participants in the asymptomatic carriers group. After applying all the filters described above, 38 phenotypes were considered for further analyses.

To evaluate the association with absence of symptoms, we compared the distribution of each variable between asymptomatic carriers and asymptomatic non-carriers. We used the lmekin() function of the R package coxme v2.2 ^43^ to fit a linear mixed model that predicts the phenotype under study based on the carrier status (outcome) and adjusted the model by age, age squared, the first four genetic PCs and sex, while accounting for familial relationships. Specifically, we used the following model:

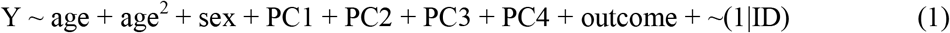

where *∼(1*|*ID)* is a random effect equivalent to twice the kinship matrix and *outcome* is a variable coded as 1 for asymptomatic carriers and 0 for asymptomatic non-carriers. Genetic PCs were included to rule out potential confounding effects of different geographic distribution of carriers and non-carriers.

### PGS definition and analysis

We downloaded summary statistics of GWAS on quantitative phenotypes and diseases related to cardiovascular function in the public repositories (Supplementary table 3). When more than one study or repository was found for the same trait, we kept the study with the largest sample size and the highest number of available SNPs. In total, we found summary statistics for a 89 traits (55 cardiac and circulatory system diseases, 10 cardiac magnetic resonance parameters, 15 echocardiographic parameters, 4 electrocardiographic parameters, 2 HR variability parameters and 3 anthropometric measures as control).

The selected summary statistics were used to calculate PGSs using the PRS-CS software, which uses the full summary statistics in a Bayesian regression framework by placing a continuous shrinkage prior on the effect size of SNPs while adjusting by the linkage disequilibrium from a reference panel.^44^ This method has been shown to be more efficient, in multiple settings, than the classical clumping and thresholding method.^45^ We used the default parameters for all our calculations.

The resulting PGSs were used to identify the genetic signatures of the phenotypes they represent and to assess if these signatures are associated with absence of symptoms in PLN c.40_42delAGA carriers. We compared the distribution of PGSs between asymptomatic carriers and asymptomatic non-carriers using the following linear model:

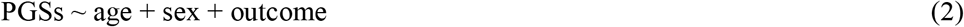

where *outcome* is a variable coded as 1 for asymptomatic carriers and 0 for asymptomatic non-carriers.

PGSs in this model were used as continuous percentiles of the standardized PGS. We also analyzed the effect of having an extreme PGS value by recoding the PGS variable as binary in the model. Specifically, we assessed the impact of having very high PGS by recoding the variable to 1 when PGS values were above the 80^th^ percentile and to 0 otherwise. Likewise, we assessed the impact of low PGS values by recoding the variable to 1 when PGS values were below the 20^th^ percentile and to 0 otherwise.

### Genetic association analyses

To evaluate the genomic region nearby the PLN c.40_42delAGA variant, we used the imputed genotypes of all genetic variants located within ±2Mb with an imputation information score higher than 0.8 and with no Mendelian errors and we phased the entire Lifelines cohort using SHAPEITv2.904^46^ with the options *–duohmm* and *–output-max*. Focusing only on the 74 carriers and the phased chromosome carrying the variant, we screened the haplotypes in increasing windows surrounding the PLN pArg14del variant. We started with the PLN pArg14del variant and added one SNP on each side to form a window and counted how many different haplotype-alleles were present among carriers. We then repeatedly added one SNP to each side. If the addition of a SNP to either side resulted in a split of the most frequent haplotype of more than a threshold number of samples (a split threshold), we stopped adding SNPs to this side and continued the other. To identify the longest shared haplotype among carriers of the PLN pArg14del variant, we first selected SNPs with MAF ≥ 0.01 and set the split threshold at 3. To include haplotypes that could possibly explain the difference in symptoms, we selected SNPs with minor allele count ≥ 4 in the group of carriers and set the split threshold at 10. We then compared the frequency of the most frequent haplotypes between symptomatic and asymptomatic carriers suing a chi-square test, only in the main splits (split threshold ≥ 3). We used the *Ghap* version 2.0 package in R^47^ for the statistical analyses and counting of the haplotypes.

To assess the potential modifier effect of common variants, we carried out a genome-wide association scan and compared allele-dosages of each variant between asymptomatic carriers and asymptomatic non-carriers using plink2.^48^ For this analysis, we only used unrelated samples because the currently available software that can account for family-based cohorts cannot properly analyze such an imbalanced number of cases (asymptomatic carriers) and controls (asymptomatic non-carriers). To identify unrelated samples, we first calculated pairwise kinship coefficients using KING^49^ v2.2 and removed individuals with high kinship (>0.125) in their respective group. For each pair of related individuals, we removed the individual of lower age. We also removed individuals in the group of controls who were related to individuals in the cases group. In all, we investigated the additive effects of 16,478,162 common variants (after filtering by MAF>0.1 for asymptomatic carriers and MAF>0.05 for non-carriers) with a logistic model that included the first PC, age, gender and the dosages of imputed or genotyped genetic variants, as in the following model:

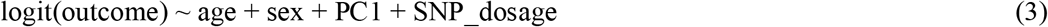

To screen for association signals that could be driven by an imbalanced allocation of variants in the long haplotype of the c.40_42delAGA variant, and are thus not related to the absence of symptoms, we assessed the correlation between the strength of the association (p-value) of SNPs with p-value ≤ 5×10^-4^ and their linkage disequilibrium (r2) with the PLN c.40_42delAGA variant. SNPs not fitting on the line of this correlation (by visual inspection) were considered suggestive associations and further investigated (**Supplementary Figure 14**).

To evaluate the effect of rare variants, we carried out region-based association tests only on low frequency (MAF ≤ 0.05) or rare (MAF ≤ 0.01) SNPs using the robust omnibus approach in the package SKAT.^50^ This approach combines variance component tests and variant collapsing tests and corrects for case–control imbalance in binary traits.^39^ We interrogated the regions ±50Kb in and around 80 genes with known genetic associations to cardiovascular diseases and ECG measurements^51^ and regions ±250Kb around the two variants identified by our GWAS. For these analyses, we used the same logistic model and samples as in equation 3. We confirmed our results using dosages and adjusting for the common variant effect. For the significant genes and regions (p<2.5×10^-6^),^52^ we further analyzed the subregion level by splitting the significant region into ten segments.

### Transcriptome-wide association study

We performed a TWAS using PrediXcan.^26,53,54^ We used the algorithm of PrediXcan to predict individual gene expression levels based on their genotypes. The models, available from PredictDB (http://predictdb.org), are elastic net–based models trained on European populations in GTex version 8 release (elastic_net_eqtl.tar). For four heart-related tissues (artery aorta, artery tibial, heart atrial appendage and heart left ventricle), we predicted expression profiles for all the individuals included in our study (N=36,339). Specifically, we imputed gene expression for 7,589 genes in artery aorta, 8,610 genes in artery tibial, 6,632 genes in heart atrial appendage and 6,006 genes in heart left ventricle in all Lifelines participants. We then performed logistic regression to investigate differences in gene expression between asymptomatic carriers and asymptomatic non-carriers, with age, gender and the first 4 PCs from the genotype data as covariates. Specifically, we used the following model:

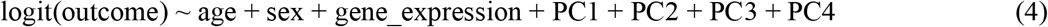

Further, we adjusted the p-values with Bonferroni correction (implemented with the Statsmodels Python package^55^ using the statsmodels.stats.multitest.multipletests() function, with the method variable being ‘bonferroni’). Similar to our GWAS, we used the same model (4) to investigate the differences for the suggestive genes in the remaining groups comparisons.

### Validation in the additional Lifelines subset cohort

We first sought to replicate our results in a second subset of 21,771 Lifelines participants for whom the same phenotype variables were available. This subset included individuals who were not genotyped with the Infinium Global Screening array but rather with the FinnGen ThermoFisher Axiom custom Array (See **URLs**), which also includes the PLN c.40_42delAGA variant. As the dataset had been recently genotyped, no imputed data was available. Neither of the two genetic variants showing suggestive association (SNP rs6768326 on chromosome 3 and SNP rs112525682 on chromosome 16) were directly genotyped with the Axiom Array. For analyses, we classified subjects to be asymptomatic or symptomatic carriers or non-carriers following the same definitions we used for our discovery dataset and used the same models described above (models 1 and 3, with the exception of principal components as they were unavailable in the second subset) to investigate the effect of quantitative phenotypes and genetic variants on asymptomatic carriers. To properly account for known relationships between the discovery Lifelines dataset and this second subset, we also jointly analyzed all Lifelines samples while correcting for the pedigree-derived kinship matrix and the subset.

### Validation in the ACM/PLN patient registry

We sought to further validate the HR and QRS phenotypes as potentially protective factors in 592 PLN c.40_42delAGA carriers from the Netherlands Arrhythmogenic Cardiomyopathy (ACM) Registry^22^, a national observational cohort study that includes patients with a definite ACM diagnosis and their at-risk relatives. The Registry is coordinated by the Netherlands Heart Institute (NHI, Utrecht, The Netherlands) and follows the Code of Conduct and the Use of Data in Heath Research and the national inclusion of patients is exempt from the Medical Research Involving Human Subjects Act (WMO) as per judgement of the Medical Ethics Committee (METC 18-126/C, Utrecht, The Netherlands). The ACM/PLN Registry is registered at the Netherlands Trial Registry, project 7097 (see **URLs**).

The registry collects participants’ medical history, and (non-)invasive test information (e.g. electrocardiograms, Holter recordings, imaging and electrophysiological studies, pathology reports, etc.) at baseline and follow-up visits. No genome-wide array data was available in the registry. For analyses, we classified subjects as asymptomatic or symptomatic carriers or non-carriers according to the definition used in our discovery dataset and used the same models as described above (models 1 and 3) to investigate the effect of quantitative phenotypes on asymptomatic carriers. Out of the 947 individuals in the registry, we excluded individuals who could not be classified as ‘asymptomatic’ or ‘symptomatic’ because of missing values, leaving 592 individuals for further analysis. We further evaluated if the observed associations remained significant and showed consistent effect directions when defining the symptomatic and asymptomatic status using additional outcome parameters collected in the ACM/PLN registry^22^, including (non)sustained VA, intracardiac defibrillator interventions, atrial arrhythmias, HF symptoms, hospitalizations and (cardiac) death (**Supplementary Table 10**). Additionally, to eliminate possible false correlations introduced by age differences, we further stratified the participants into younger (≤40 years of age) and older (>40 years) groups and repeated the regression analysis. For the analyses in the ACM/PLN registry, we did not remove individuals from the same family because the exact family relationships were not recorded.

We tested the predictability of QRS duration in predicting the VA risk for PLN c.40_42delAGA carriers. For this, we first correlated QRS duration with the predictors used in the model from Verstraelen et al.^13^: the number of negative T waves, presence of low-voltage ECG, PVC count/24 h and LVEF. Based on the correlation results, we identified two predictors that are highly correlated with QRS duration: PVC count/24h and LVEF. We then tested whether QRS duration could partly reflect the predictability for those predictors by comparing the prediction performance between two models: 1) a baseline model that excluded the highly correlated predictors with QRS duration and 2) a model with the predictors used in model (1) and QRS duration. We used leave-one-out AUC value to quantify the prediction performance and determined the significance of the differences between the two models using the DeLong test.^56^ Model construction and evaluation were implemented using Python package scikit-learn v0.22.2.post1^57^ and R package pROC v1.14.0.^56^

### Data availability Statement

Genotyping data and participant metadata are not publicly available to protect participants’ privacy and cannot be deposited in public repositories to respect the research agreements in the informed consent. Data can be accessed by all bona-fide researchers with a scientific proposal by contacting the Lifelines Biobank (https://www.lifelines.nl/researcher/how-to-apply). Researchers will need to fill in an application form that will be reviewed within 2 weeks. If the proposed research complies with Lifelines regulations, such as noncommercial use and guarantee of participants’ privacy, applying researchers will receive a financial offer and a data and a material transfer agreement to sign. In general, data will be released within 2 weeks after signing the offer and the data and material transfer agreement. The data will be released in a remote system (the Lifelines workspace) running on a high-performance computer cluster to ensure data quality and security. The ACM/PLN registry data can be requested in writing to. All summary data supporting the conclusions of this data are given in Supplementary Tables.

## Supplementary information

See supplementary information and supplementary tables at the publication site.

## URLs

Lifelines catalog: https://data-catalogue.lifelines.nl/@molgenis-experimental/molgenis-app-Lifelines-webshop/dist/index.html#/shop/

ACM/PLN Registry:

https://www.acmregistry.nl/

https://www.trialregister.nl/trial/6902

FInnGen thermoFisher Axiom custom Array:

https://www.finngen.fi/en/researchers/genotyping

