## Supplementary information for "Phenotypic and genetic factors are associated with absence of cardiomyopathy symptoms in PLN c.40_42delAGA carriers" (from another version of this preprint: version 1, 2022-05-16)

**Supplementary Tables**

S1. Phenotype description

S2. Phenotype associations

S3. Replication of phenotypes

S4. Haplotype analyses

S5. GWAS result

S6. Rare variant analyses

S7. PGS Analysis

S8. Interaction analyses

S9. TWAS

S10. ACM symptoms

S11. City index and exercises in QRS

**Supplementary Figures**

SF1. Distribution of age in PLN p.Arg14del carriers and non-carriers

SF2. Clustering of symptomatic and asymptomatic carriers in families

SF3. Phenotype distributions according to carrier and symptoms status

SF4. Forest plot depicting the effect of PGSs

SF5. Distribution of polygenic scores compared between the four study groups.

SF6. Distribution of the symptoms in the interaction of PGS_QRS_ with the symptomatic carrier status

SF7. Differences in number of variants between the haplotypes present within the window of the longest and most frequent haplotype for the 74 PLN-p.Arg14del carriers in two screening frames.

SF8. Manhattan and regional association plots for genetic analyses between asymptomatic carriers compared to asymptomatic non-carriers

SF9. TWAS results in the Heart Left Ventricle

SF10. Phenotype distributions for symptomatic and asymptomatic individuals in the ACM registry

SF11. Correlation between MRI-derived features and ECG features from Verstraelen et al. study and QRS durations in the ACM registry.

SF12. Comparison of the ventricular arrhythmia risk prediction

SF13. Intensity readings of the genotype calls for the Illumina variant seq-rs397516784 (corresponding to PLNp.Arg14del)

SF14. Correlations between the variant’s GWAS p-value and its linkage disequilibrium (r^2^) to PLN-p.Arg14del for the variants with p-values < 5x10^-4^

**Supplementary Text**

**Binary analyses of polygenic scores (PGSs)**

As we were concerned that PGSs could have some non-linear effects, we repeated our analyses of PGSs, but focusing on the extremes of the distributions. Specifically, we compared the number of asymptomatic carriers and asymptomatic non-carriers in the upper tail (PGS above the 80^th^ percentile) with those in the rest of the distribution. Likewise, we compared the number of asymptomatic carriers and asymptomatic non-carriers in the lower tail (PGS below the 20^th^ percentile) against the rest. The results for these analyses were consistent with those observed when analyzing the PGS as continuous distribution only for the MRI-lvesv traits (**Supplementary Tables 7D and E**).

**Sensitivity analyses of the significant common variants on chromosome 6 identified by GWAS**

We sought to rule out any potential confounding effects due to the fact that PLN Arg14del variant carriers have a long haplotype that includes common variants that would then be differentially distributed between carriers and non-carriers, regardless of symptoms. The top association on chromosome 6 was observed with SNP rs78559806, which is in low LD (r^2^=0.01, D’=0.83) with the PLN-Arg14del variant. When correlating the strength of the associations observed across chromosome 6 (GWAS p-values) with the LD (r^2^) to PLN-del variant, we found a significant correlation (rho=0.92, p<2.2x10^-16^), indicating that the stretch of associations on chromosome 6 is not related to lack or presence of symptoms (**Supplementary Figure 14**).

**Independent effect of QRS and PGS_QRS_ on asymptomatic carriers compared to asymptomatic non-carriers**

We also found a higher predisposition for increased PGS_QRS_ in carriers, which may appear to be in contrast with decreased QRS we observed in asymptomatic carriers. Considering that PGS_QRS_ showed (as expected) a positive correlation with QRS duration (Spearman correlation rho=0.18, p=1.42x10^-182^, using all participants), we hypothesized that the effects of the QRS phenotype and the PGS_QRS_ were independent. To test this hypothesis, we jointly analyzed the PGS_QRS_ and the phenotypic value of QRS duration in one model. Both terms remained significant, and the effect of one was not diminished by the presence of the other in the model (**Supplementary Table 8**), which corroborates that the effects of PGS_QRS_ and QRS on our outcome were truly opposite and independent.

**Associations of QRS duration with participant physical activity measures and urban index of their residence**

We collected residential information for the Lifelines participants and mapped their urban index using information from a previous study^57^. We then examined the distribution of the urban index for the four groups (asymptomatic carriers, asymptomatic non-carriers, symptomatic carriers, and symptomatic non-carriers) of participants in Lifelines with the following model and OLS:

QRS duration ~ group + urban1+ age + gender + group * urban index

The *group* variable in the model indicates whether a participant is a carrier and whether a participant is symptomatic. The results of the model showed no significant findings for any variable related to the urban index (**Supplementary Table 11**).

We also collected participant lifestyle information about sports frequency, sedentary duration, exercise duration last week and regular sports duration. The specific parameter names are:

'exercise_pastweek_ach_q_1_1A', 'sports_frequency_ach_q_1_01', 'sports_frequency_ach_q_1_02', 'sports_frequency_ach_q_1_03', 'sports_play_ach_q_1_01', 'sports_play_ach_q_1_02', 'sports_play_ach_q_1_03', 'sports_regular_ach_q_1', 'activity_followup_adu_q_1.B', 'activity_followup_adu_q_1.C', 'exercise_pastweek_ach_q_1_2A', 'sedentary_duration_ach_q_1', 'sport1_hours_ach_q_1', 'sport2_hours_ach_q_1', 'sport3_hours_ach_q_1'.

The explanation for each parameter name can be found in the Lifelines catalog website (See **URL section**). We then assessed the correlation between these exercise variables with the participant's QRS duration values using an OLS model adjusted for gender and age but did not find significant associations between any exercise variables and QRS duration (**Supplementary Table 8**).

**Supplementary Figures**

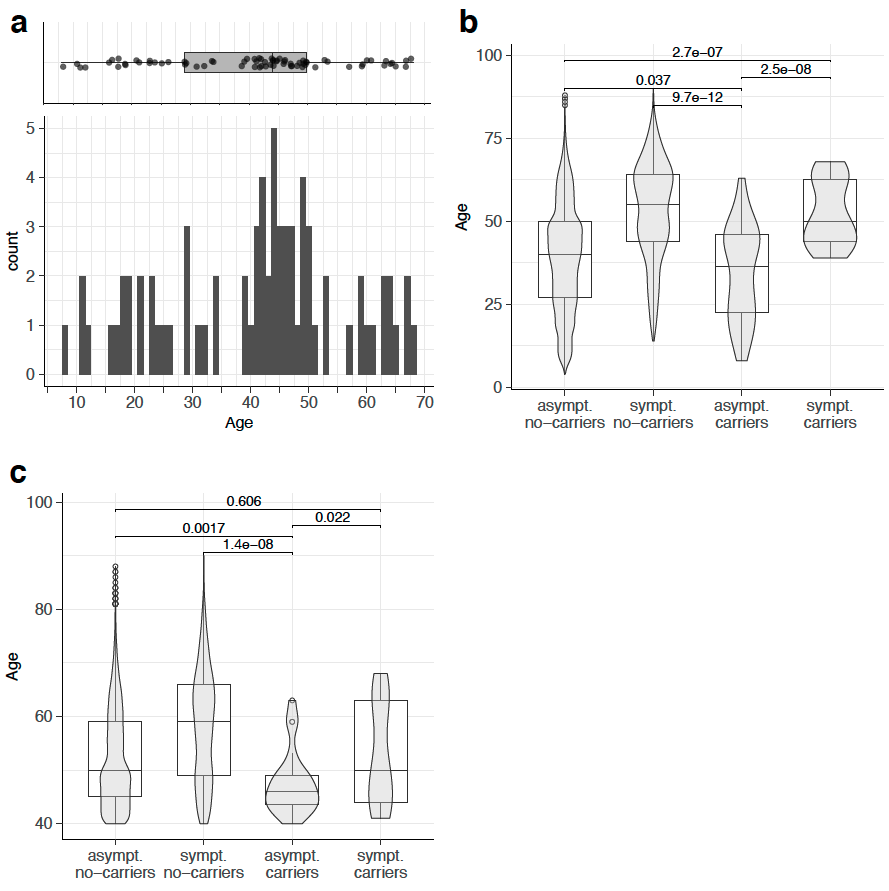

**Supplementary Figure 1.** **Distribution of age in PLN p.Arg14del carriers and non-carriers.** a) Boxplot of age distribution (top) and frequency by age (bottom) for the 74 carriers only. b) Boxplot and violin plots for age in each of the four groups: asymptomatic non-carriers (n=34,201), symptomatic non-carriers (n=2,064), asymptomatic carriers (n=48) and symptomatic carriers (n=26). c) Box plots and violin plots for age when including only individuals aged ≥40 years in each of the four groups, asymptomatic non-carriers (n=17,554), symptomatic non-carriers (n=1,705), asymptomatic carriers (n=23) and symptomatic carriers (n=25). *p-values correspond to pairwise t-tests.

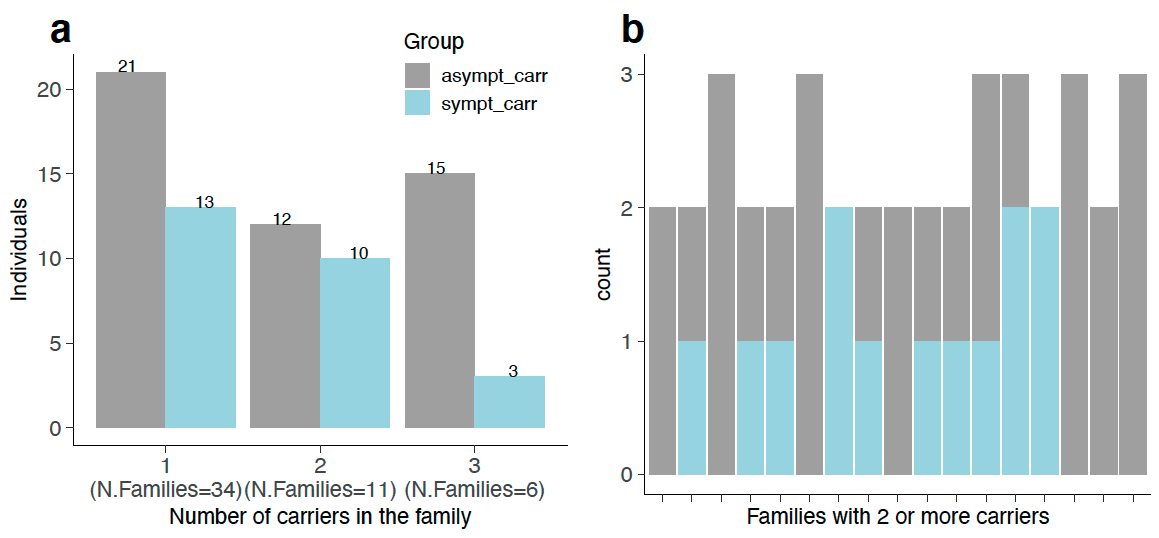

**Supplementary Figure 2. Clustering of symptomatic and asymptomatic carriers in families.** a) Number of symptomatic and asymptomatic individuals per family, with the families represented in the X-axis by the number of carriers they contain. b) Numbers of carriers in the 17 families with more than one carrier. Each frequency bar is one family, and the segments represent the individual carriers, colored according to the presence or absence of symptoms. Only seven symptomatic carriers are in the same family as asymptomatic carriers. asympt_carr. asymptomatic carriers; sympt_carr, symptomatic carriers.

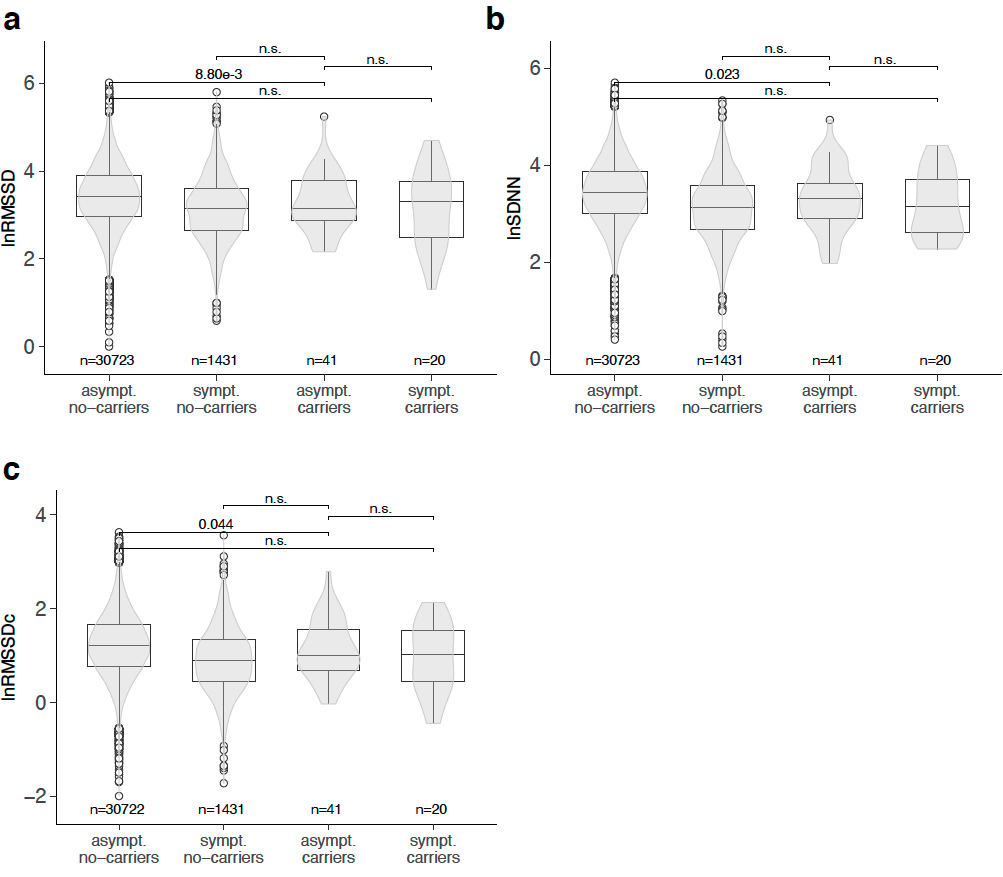

**Supplementary Figure 3. Phenotype distributions according to carrier and symptoms status.** Boxplots and violin plots represent quantitative phenotypes as described in **Figure 1.** a) Boxplot and violin plots for adjusted root mean square of successive differences adjusted by heart rate. b) Boxplot and violin plots for root mean square of successive differences. c) Boxplot and violin plots for the standard deviation of normal-to-normal intervals. The symptomatic carriers group shows decreased values in all three heart rate variability measures that appear to be unique to this group. This difference is apparently not affected by heart rate itself, as shown in (c). p-values correspond to the coefficient of the adjusted trait in the model described in Equation 1.

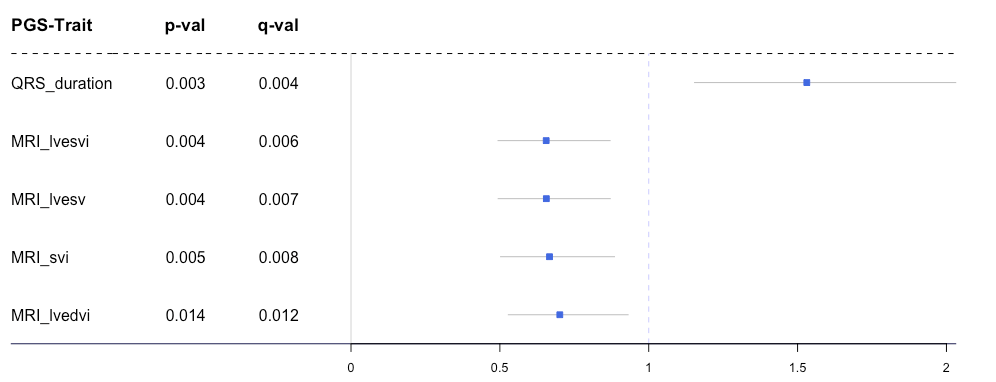

**Supplementary Figure 4**. **Forest plot depicting the effect of PGSs.** Squares represent the effect size (beta). Lines represent standard error. p-val, p-value for the independent association; q-val, false discovery rate for conjoined analysis of all the traits, with 1000 permutations for each of the 97 PGS-traits.

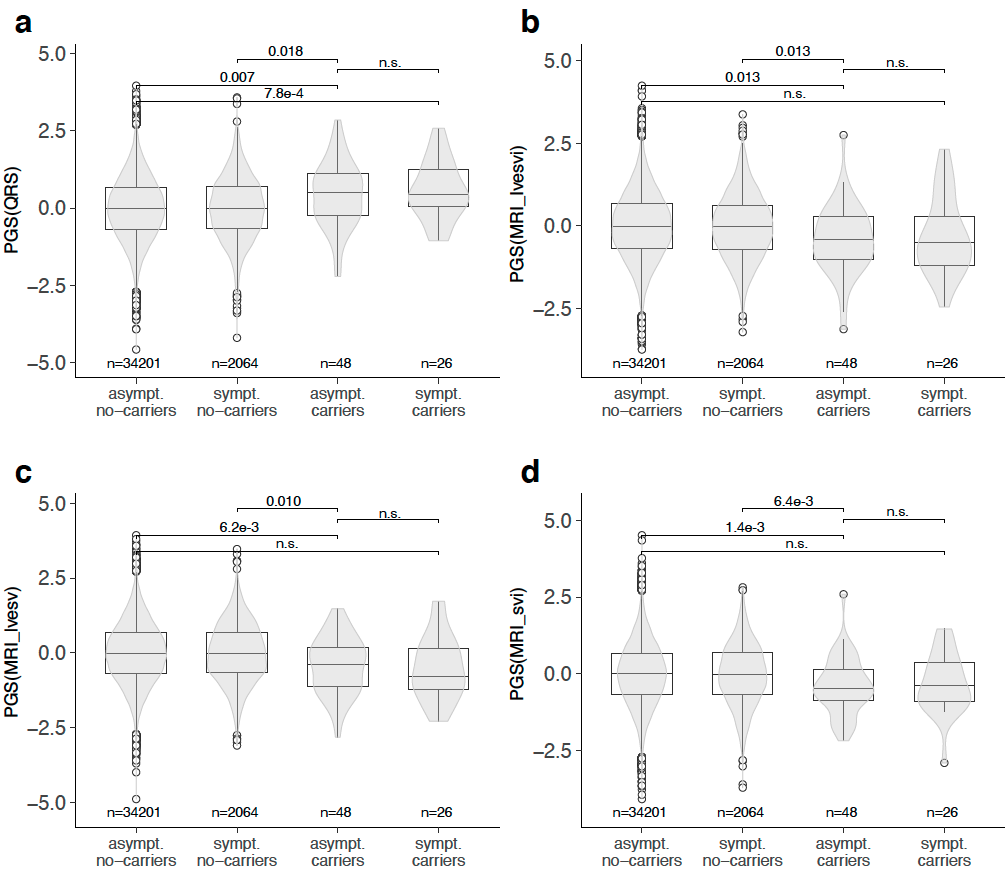

**Supplementary Figure 5.** **Distribution of polygenic scores in the four study groups.** Violin and boxplots represent each PGS as described in **Figure 1**. Comparisons are shown for a) PGS_QRS_, b) PGS_lvesvi_, c) PGS_lvesv_ and d) PGS_svi_. The distribution of asymptomatic carriers is not different from that of the symptomatic carriers, while the difference compared to the symptomatic non-carriers for both groups of carriers has the same direction. P-values correspond to the regression coefficient of the adjusted trait in the model described in equation 2, comparing the groups indicated by the ticks. asympt, asymptomatic; sympt, symptomatic.

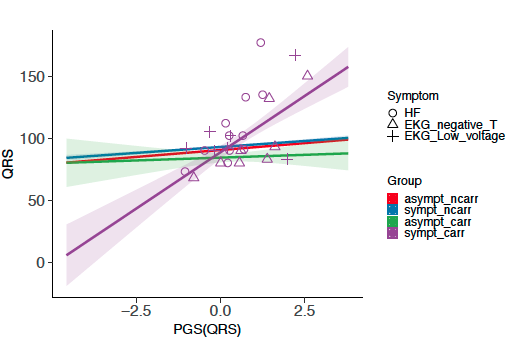

**Supplementary Figure 6. Distribution of symptoms in the interaction of PGS_QRS_ with the symptomatic carrier status.** Regression lines represent the partial correlation between PGS_QRS_ and QRS for each of the groups. Symbols represent the symptoms only for symptomatic carriers, ordered according to severity as in the legend from highest to lowest. For individuals with more than one symptom, only the most severe is depicted. There is no apparent pattern in the severity in this interaction. HF, heart failure, EKG_negative_T, negative T in the electrocardiogram; EKG_Low_voltage, low voltages or microvoltages in the electrocardiogram.

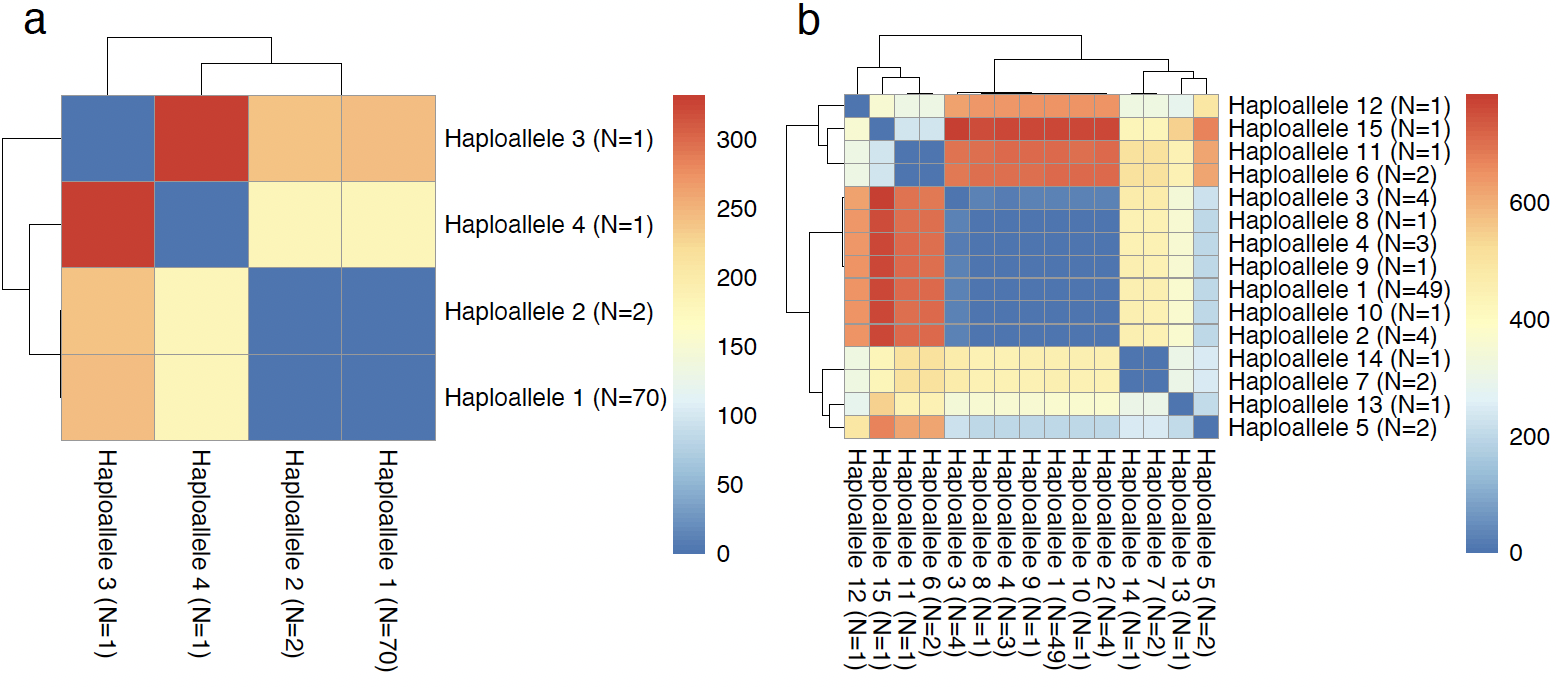

**Supplementary Figure 7. Differences in the number of variants between the haplotypes present within the window of the longest most frequent haplotype for the 74 PLN-p.Arg14del carriers** **in two screening frames**. a) For a MAF ≥ 0.01 in all the samples. The window comprises 1.38Mb (+605Kb to -770Kb around PLN-p.Arg14del). Haploalelle 2 is closest to the most frequent haploallele (n=70) with 1 SNP of difference, whereas the other haploalleles differ by approximately 330 SNPs. b) For a minor allele count ≥ 4 in the carrier chromosomes. The window comprises 2.25Mb (+245Kb to -1.98Mb), and we did not stop the screening until we reached a split (at least 10 haploallele difference). Therefore, the window extended until the upstream end of the region screened. Other haploalleles differ from the longest most frequent haploallele (n=49) in a range of 2 and 780 SNPs.

**a)**

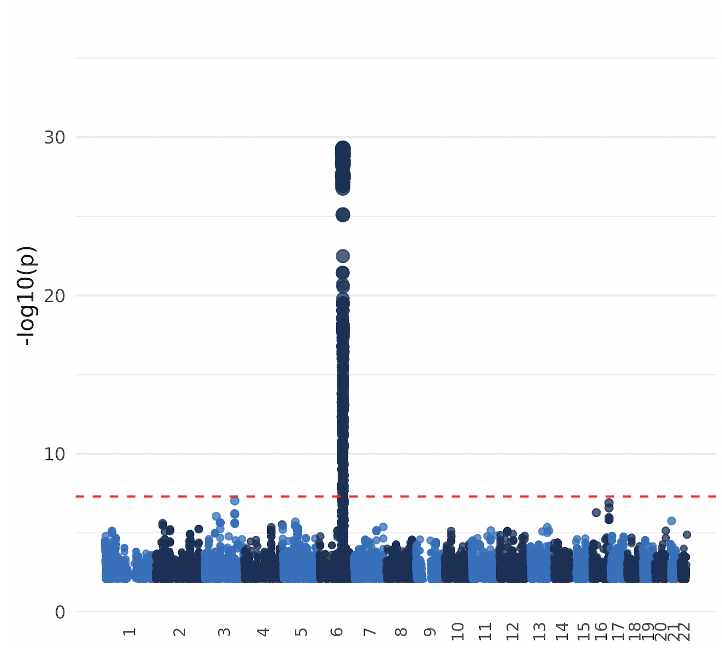

**b)**

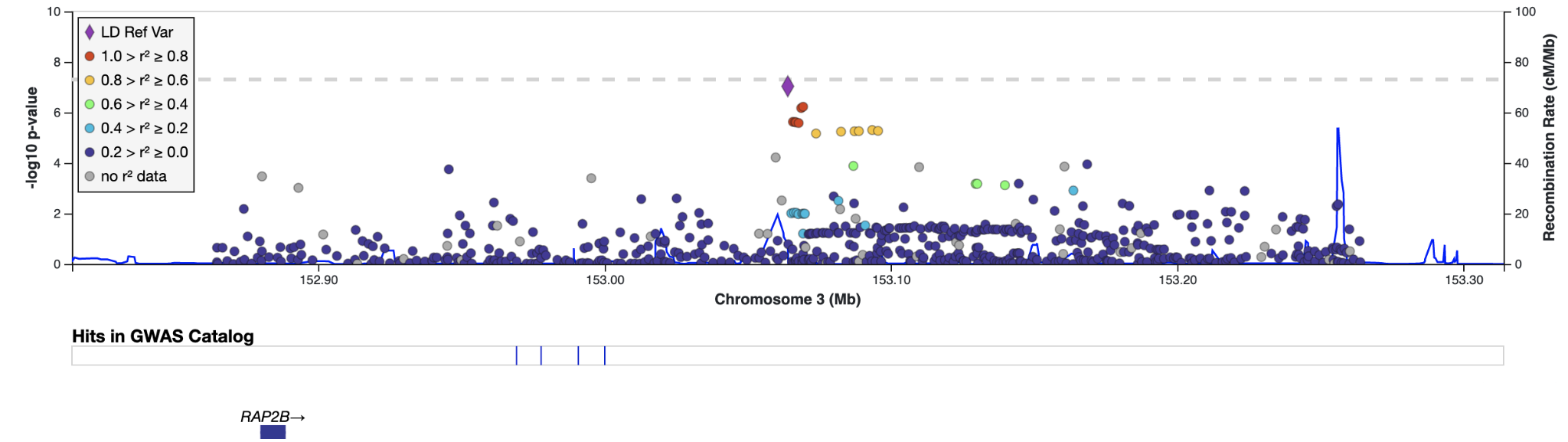

**c)**

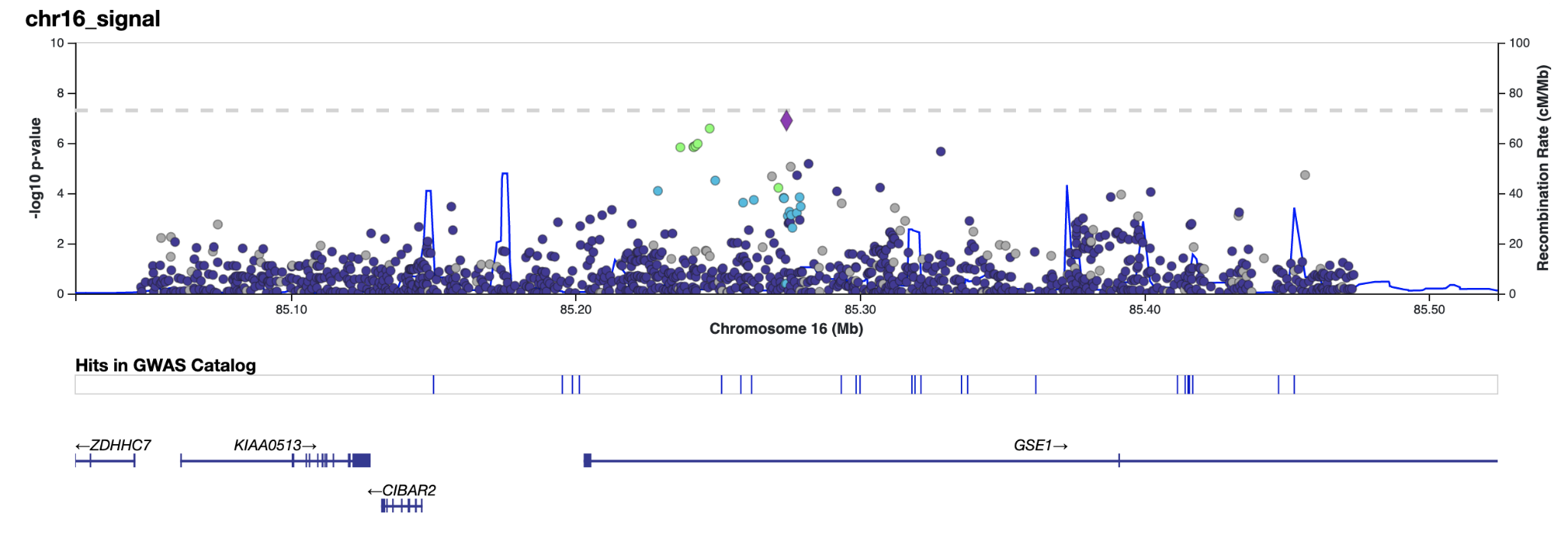

**Supplementary Figure 8. Manhattan and regional association plots for genetic analyses between asymptomatic carriers compared to asymptomatic non-carriers.** a) Genome-wide comparison. Dotted red line corresponds to the genome-wide significance threshold (p=5x10^-8^). b) Locus-zoom plot for 200Kb around rs6768326 in chromosome 3. c) Locus-zoom plot for 200Kb around rs112525682 in chromosome 16. LD measures correspond to EUR population.

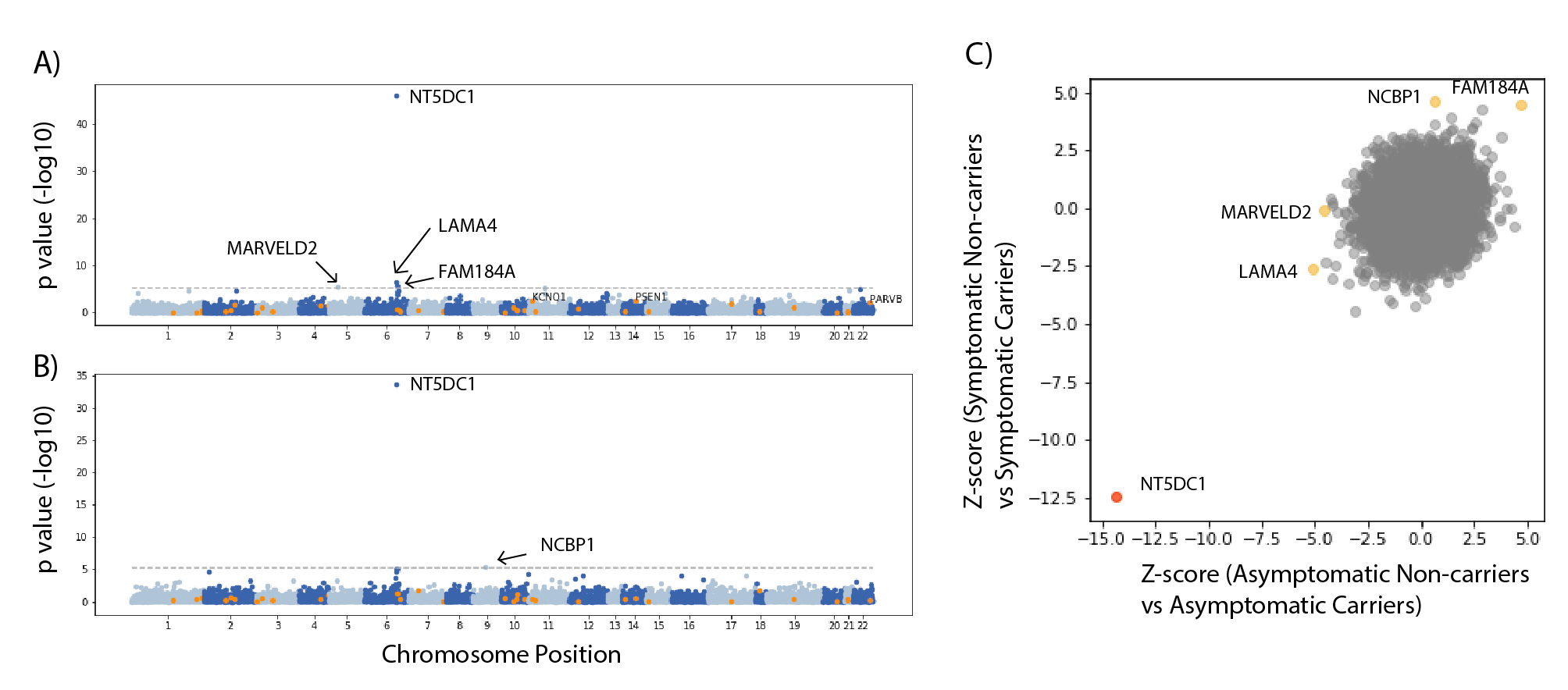

**Supplementary Figure 9**. **TWAS results in Heart Left Ventricle.** a) For asymptomatic carriers compared to asymptomatic non-carriers. b) For symptomatic carriers compared to symptomatic non-carriers. Significance level is determined as -log(p) < 5.18. Each dot in the figure represents a gene. Orange dots are genes in the cardiomyopathy panel. c**)** Comparison between the z-scores of TWAS results for asymptomatic carriers and those for symptomatic carriers. Dark orange dots indicate genes significant in both TWAS studies. Light orange dots indicate genes significant in only one TWAS study.

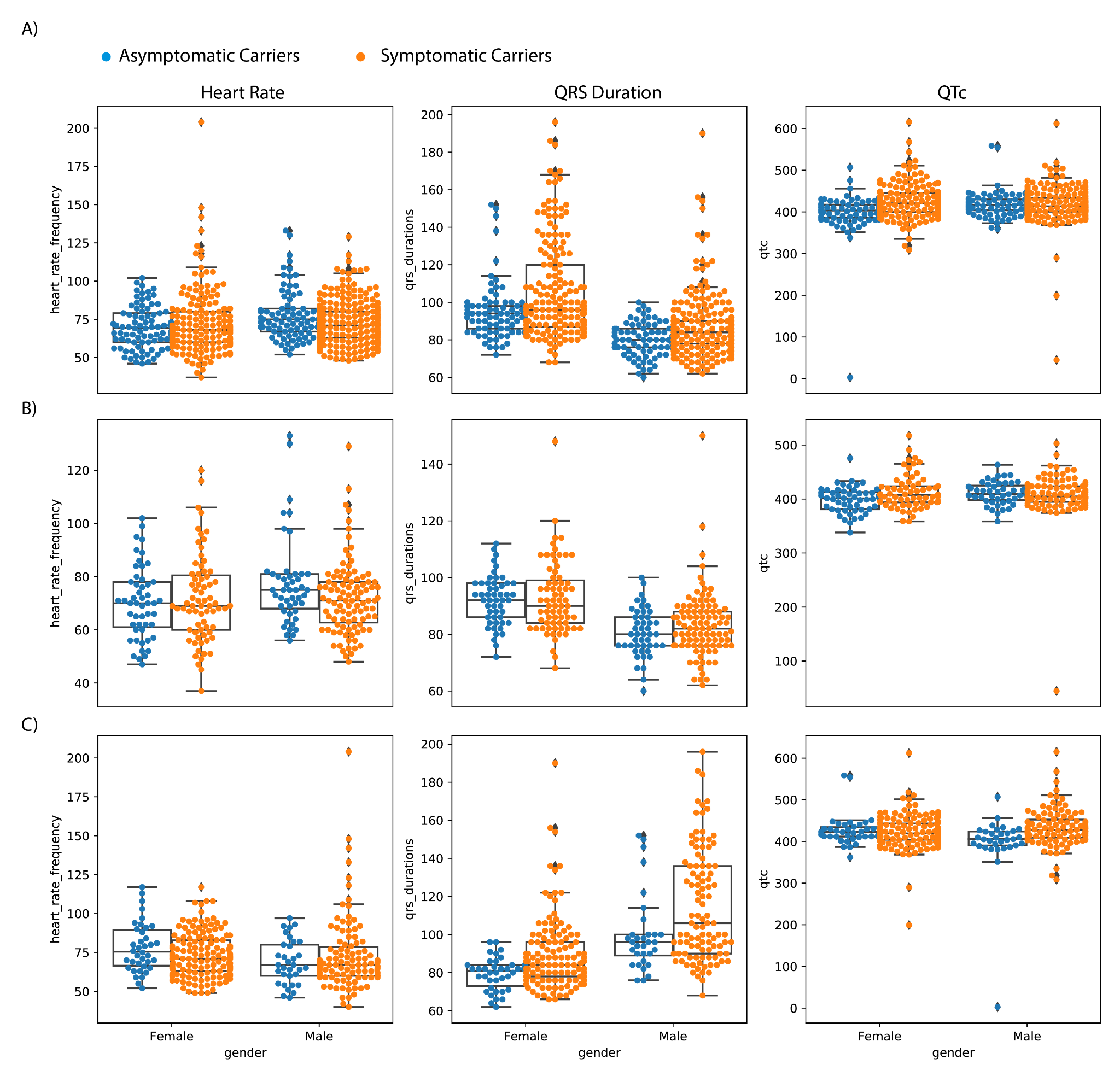

**Supplementary Figure 10.** **Phenotype distributions for symptomatic and asymptomatic individuals in the ACM registry.** a) Phenotype distribution for the full ACM registry data. b) Phenotype distribution for younger people (age < 40 years) in the ACM registry. c) Phenotype distribution for old people (age ≥ 40 years) in the ACM registry.

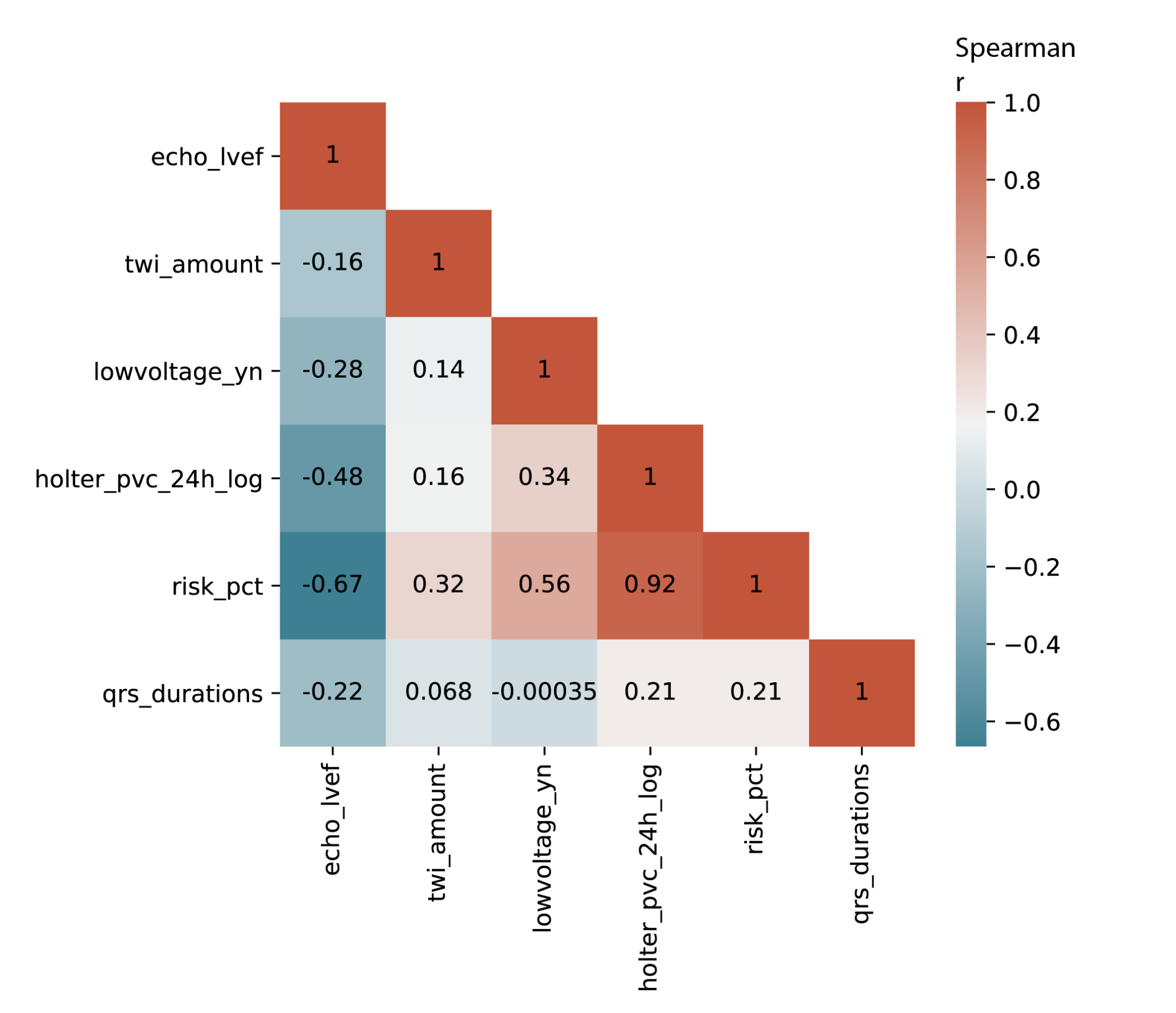

**Supplementary Figure 11.** **Correlation between MRI-derived features and ECG features from the Verstraelen et al. study and QRS durations in the ACM registry.** Box color indicates Spearman r value.

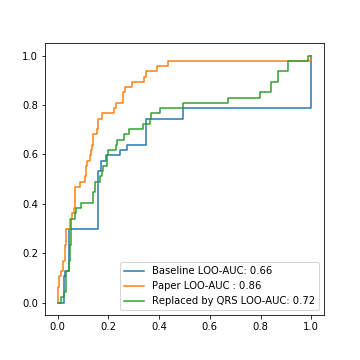

**Supplementary Figure 12**. **Comparison of the ventricular arrhythmia risk prediction.** “Paper” indicates that this model is obtained using the same set of features used in the Verstraelen et al. study: LVEF, 24-h PVC counts, T wave inversion and the presence of low voltage ECG. “Replaced by QRS” indicates that the model is obtained using QRS duration, T wave inversion and the presence of low voltage ECG. “Baseline” indicates that this model is obtained using only T wave inversion and the presence of low voltage ECG.

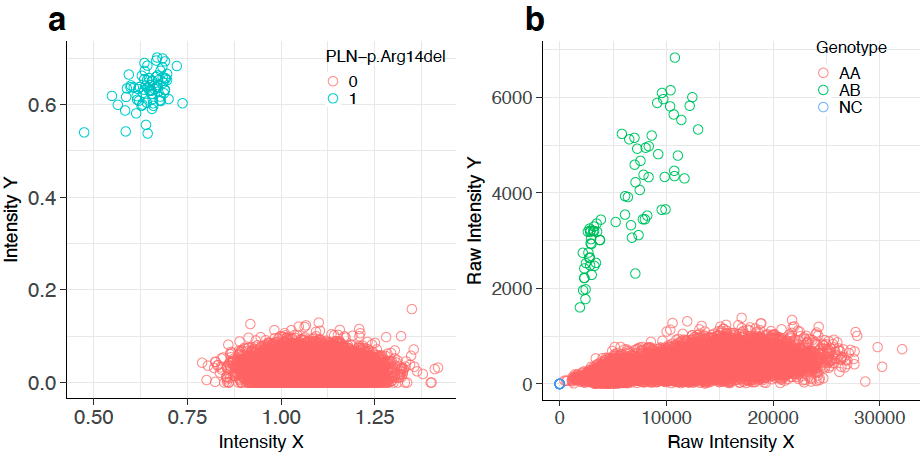

**Supplementary Figure 13.** **Intensity readings of the genotype calls for the Illumina variant seq-rs397516784 (corresponding to PLNp.Arg14del).** a) Genotype calls from the Illumina software with normalized intensities for the SNP. The 74 samples with high intensity values would be interpreted as carriers of alternative alleles for this SNP. b) Raw intensity readings show higher Y values for 74 samples, which can be interpreted as the heterozygous genotype (AB). These 74 samples cluster away from those homozygous for the reference allele (AA), which show low intensity values in Y and spread intensities in X. Non-calls (NC) were observed in 14 samples.

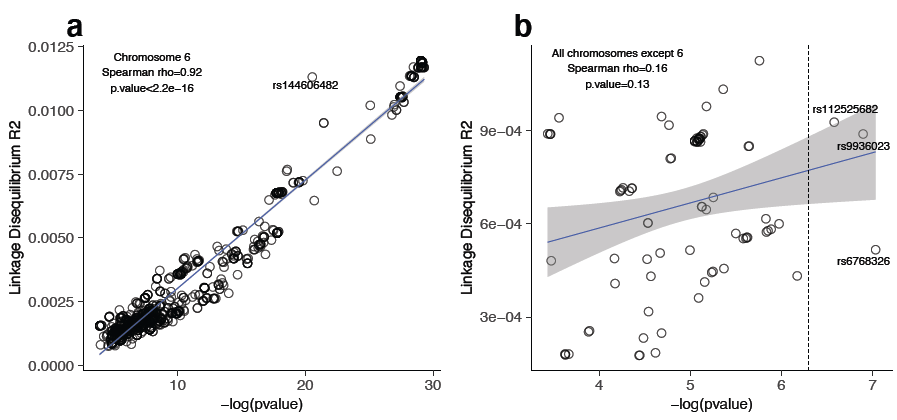

**Supplementary Figure 14. Correlations between the variant’s GWAS p-value and its linkage disequilibrium (r^2^) to PLN-p.Arg14del for variants with p-values < 5x10^-4^.** Each point represents a genetic variant. a) Correlation for SNPs located in chromosome 6 only. b) Correlation with SNPs located in the rest of the genome. SNPs outside of this correlation and with a p-value < 5x10^-7^ (dashed line) were labeled and further analyzed. Regression lines are depicted in blue.
